# Impact of simulated MRI artifacts on deep learning-based brain age prediction

**DOI:** 10.64898/2026.03.24.26349152

**Authors:** Janine Hendriks, Michelle G. Jansen, Richard Joules, Óscar Peña-Nogales, Femke Elsen, Anya Povolotskaya, Mathijs B.J. Dijsselhof, Paulo R. Rodrigues, Frederik Barkhof, Anouk Schrantee, Henk J.M.M. Mutsaerts

## Abstract

Brain age is a promising biomarker for detecting atypical and pathological brain aging, but its accuracy and reliability depend critically on MRI quality. The impact of common MR image degradations such as motion, ghosting, blurring, and noise on brain age predictions remains unclear. In this study, we systematically assessed the effects of four simulated MRI artifact types, across ten severity levels, on brain age prediction using three widely used deep learning–based algorithms (Pyment, MIDI, MCCQR), in high-quality T1-weighted images of healthy adults (age range 18-85, 54% female). Artifact severity levels (1–10) were generated using a power-function mapping of TorchIO simulation parameters calibrated to the full PondrAI QC visual rating scale (from perfect to severely degraded image quality). Linear mixed-effects models with predicted brain age as dependent variable revealed a significant interaction between algorithm, artifact type, and severity (p<0.001), indicating algorithm-specific sensitivity to artifacts. In artifact-free scans, mean absolute error (MAE) was 4.6 years for MCCQR, 7.1 years for Pyment, and 9.1 years for MIDI. At severity level 10, MAE increased with up to 110% for Pyment, 112% for MCCQR, and 16% for MIDI (motion); and with up to 75% for Pyment, 135% for MCCQR, and 34% for MIDI (ghosting). Blurring had minimal impact at low–moderate levels, but at maximum severity MAE increased by 26% for Pyment and 137% for MCCQR, while MIDI remained largely stable. Noise minimally affected Pyment and MCCQR (MAE increases ≤20%), but led to larger declines for MIDI (MAE increase 35%). The vulnerability of different algorithms highlights that training data, preprocessing strategies and underlying architectures influence robustness, emphasizing that artifact sensitivity is a key consideration when interpreting brain-age as a biomarker. Our results emphasize the need for artifact-aware evaluation and mitigation strategies when algorithms such as brain age are used in clinical research.

## 1. Introduction

Structural magnetic resonance imaging (MRI) is an important tool for non-invasive investigation of brain anatomy and pathology. T1-weighted MRI scans are commonly used to segment gray matter, white matter, and cerebrospinal fluid, enabling the derivation of metrics such as cortical thickness, brain volume, and surface area. These MRI-derived brain metrics have become central for understanding typical and atypical brain development, as well as detecting early signs of neurodegenerative or psychiatric disorders (Choi et al., 2019; Molina Galindo et al., 2024; Thompson et al., 2003).

A rapidly growing application of MRI-derived metrics is brain age prediction, which employs machine learning or deep learning algorithms to estimate an individual’s biological brain age based on structural brain features(Azzam et al., 2025; Dorfel et al., 2023). The discrepancy between predicted brain age and chronological age, known as the brain-age difference or brain-age gap, is often interpreted as a marker of accelerated aging and a potential disease predictor (Franke & Gaser, 2019). Publicly available algorithms such as brainageR (Biondo et al., 2022; Clausen et al., 2022; Hobday et al., 2022), DeepBrainNet (Bashyam et al., 2020), and Pyment (Leonardsen et al., 2022) have demonstrated high accuracy and test-retest reliability in healthy populations, supporting their methodological robustness.

The process of translating raw MRI data into reliable brain age predictions with clinical utility remains complex (Gaser et al., 2024). Quality control (QC) is a critical step, as artifacts such as motion, scanner-induced distortions, or protocol inconsistencies can significantly compromise the accuracy of downstream analyses (Valdes-Hernandez et al., 2023). Recent studies highlight the promise of deep learning for brain age prediction, especially in handling image degradation and multimodal data integration (Gaser et al., 2024). Unlike traditional machine learning, where performance often depends on extensive preprocessing that is susceptible to MRI artifacts, deep learning can be trained on minimally processed MRI data, allowing training datasets to include more representative examples of real-world image variability and artifacts, which may improve robustness to acquisition-related errors when such variability is adequately represented during training (Hanson et al., 2024; Vakli et al., 2024).

Although brain age algorithms are usually trained and tested on research-grade data, which are typically of high-quality and artifact-free, clinical data exhibits significantly larger diversity including images of poorer quality and presenting with artefacts (Lei et al., 2022). This limits our understanding of how these algorithms perform in more heterogenous, real-world context with clinical-grade data. Progress in this area is further constrained by the lack of annotated datasets that represent the full spectrum of image quality encountered in clinical practice. In particular, there is a scarcity of systematically annotated scans with mild or subtle artifacts; data that would be essential for training and evaluating algorithm robust to real-world clinical data. This scarcity has so far prevented the development and retraining of brain-age algorithms on clinically representative cases. Without such datasets, algorithms cannot be effectively retrained or calibrated on representative clinical data. As a result, most users of available brain age algorithms continue to apply pretrained algorithms originally optimized for research-quality scans, rather than retraining them on or adjusting to their own clinical data. This has important implications for translation into clinical contexts, as algorithms trained on research-grade datasets may be directly applied to clinical-grade images and perform poorly or unexpectedly. Additionally, algorithms differ in architecture and baseline prediction accuracy, as such their sensitivity to image quality and specific artifact types can also vary. Consequently, comparing differently trained algorithms (e.g., research versus clinical datasets) becomes confounded not only by differences in image quality but also by the algorithms themselves.

This study systematically assesses how different simulated MRI artifacts (motion, ghosting, blurring, and noise), representing the most frequently encountered degradations in T1-weighted structural MRI scans (Krupa & Bekiesińska-Figatowska, 2015), affect deep learning–based structural brain age prediction. Specifically, we evaluate how increasing artifact severity alters (1) deviations from artifact-free brain-age predictions, (2) prediction performance – the accuracy of predicted brain age relative to chronological age, and (3) prediction stability – the consistency of within-subject brain age predictions. We compare three widely used brain age prediction algorithms, which were pre-trained on either research-grade data or clinical-grade datasets.

## 2. Methods

### 2.1. Participants

Data were drawn from the Advanced Brain Imaging on ageing and Memory (ABRIM) dataset (Jansen et al., 2024). ABRIM is an open-access, cross-sectional neuroimaging dataset designed to investigate the neuroimaging correlates of cognitive aging and memory decline, including 295 healthy adults aged 18 to 85 years. The study design and in– and exclusion criteria are provided in detail previously (Jansen et al., 2024). Briefly, we included the 293 participants (50.9 ± 17.1 years, 54% female) with available T1-weighted Magnetization Prepared – RApid Gradient Echo (MPRAGE) images, which were acquired on a 3T Siemens Magnetom Prisma System using a 32-channel receive coil (Siemens, Erlangen, Germany; TR = 2200 ms, TE = 2.64 ms, voxel size = 0.8×0.8×0.8 mm).

The ABRIM study fell under the ethics approval “Image Human Cognition” (Commissie Mensgebonden Onderzoek Arnhem-Nijmegen, 2014/288), and was approved by the Social Sciences Institutional Review Board of the Radboud University (ECSW 2017-3001-46). The study was conducted in compliance with all local procedures and applicable national legislation and in compliance with the Helsinki declaration. All participants provided written informed consent prior to participation.

### 2.2. Artifact generation

Artifact simulation at image level was performed using the open-source Python-based toolbox TorchIO (Perez-Garcia et al., 2021). To systematically assess the impact of MRI artifacts on brain age prediction, we simulated four types of image artifacts: motion, ghosting, blurring, and noise. The original T1-weighted images were visually inspected and confirmed to be free of visible artifacts (severity level 0). We defined ten severity levels for each artifact type. The TorchIO artifact simulation parameter values and range (**Suppl. Table 1)** were based on visual quality ratings (see **Supplemental methods**). In short, these visual ratings were performed using the PondrAI QC visual rating scale, which has previously been applied to several publicly available neuroimaging cohorts (Costantino & Devenyi, 2023). PondrAI QC ranges from 1 to 6: 1 = perfect; 2 = very good; 3 = good; 4 = bad; 5 = very bad; 6 = terrible. For each artifact type, we used the visual QC ratings to guide TorchIO parameter values and ensure that the simulated artifact severity levels reflect the full visual rating scale (PondrAI QC score 1-6). To allow for a more detailed analysis of subtle degradations, we used a power function with smaller step-size for low severities (PondrAI QC scores 1-3) and larger step-size for high severities. The resulting severity levels 1 to 7 approximately correspond to images rated as “perfect” to “good” (PondrAI QC 1–3), and severity levels 8 to 10 to “bad” to “terrible” images PondrAI QC 4-6) (see **Figure 1**). For each of the four artifact types, this resulted in 293 non-degraded and 2930 degraded images, n=3223 in total.

**Figure 1.**
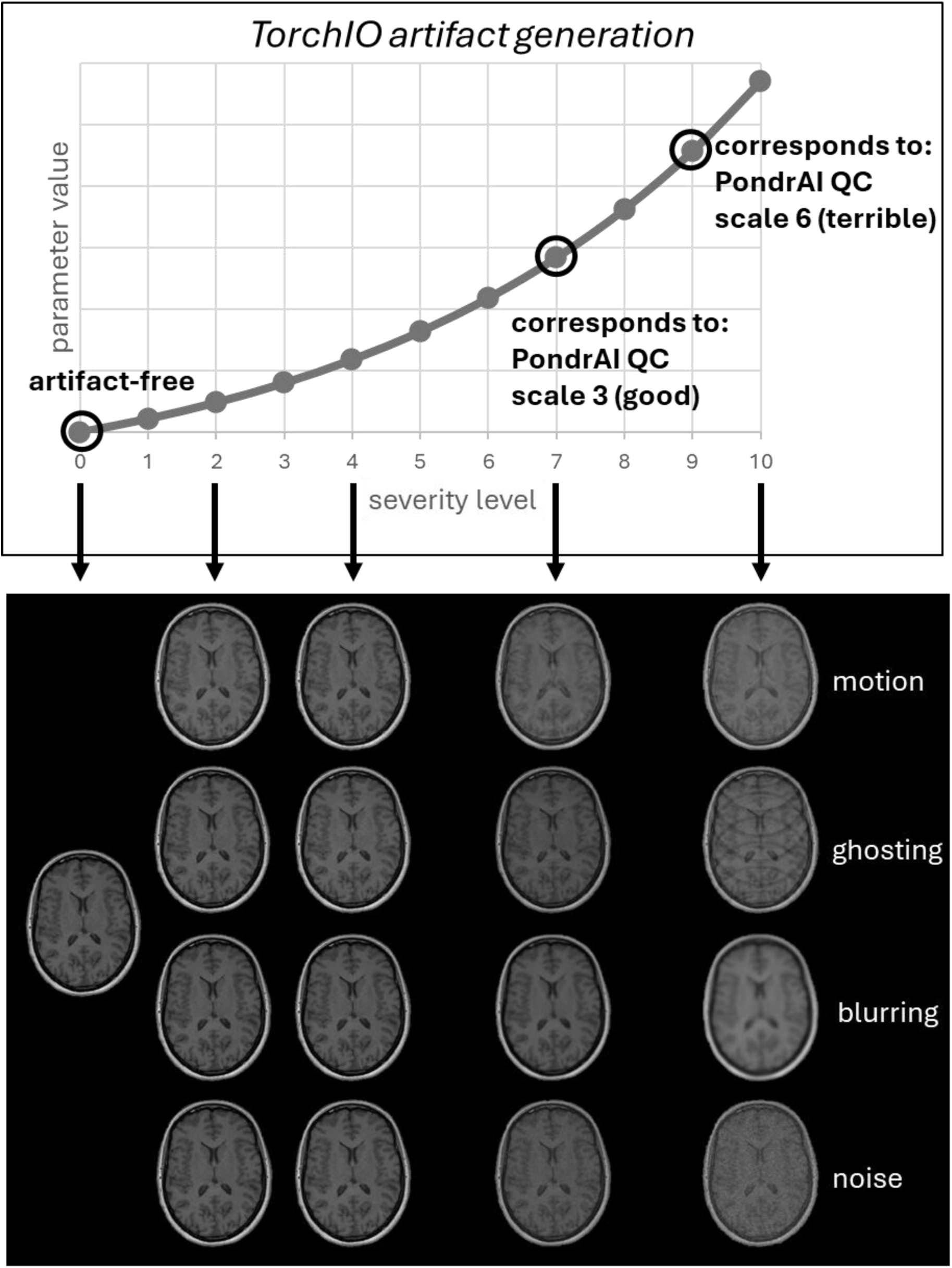
Overview of the artifact simulation at image level. Top: correspondence of artifact severity levels (x-axis) with PondrAI QC scale (y-axis). Bottom: images of a representative subject at severity level 0 (artifact-free), severity level 2, severity level 4, severity level 7 and severity level 10. PondrAI QC ranges from 1 to 6: 1 = perfect; 2 = very good; 3 = good; 4 = bad; 5 = very bad; 6 = terrible.

### 2.3. Brain age prediction algorithms

For brain age estimation, we selected three deep-learning based brain age prediction algorithms that are widely used in the literature and are trained on large datasets with T1-weighted images: Pyment (Leonardsen et al., 2022), MIDI (Wood et al., 2022), and Monte Carlo dropout Composite Quantile Regression (MCCQR) (Hahn et al., 2022) (**Figure 1B**). In their respective studies, the algorithms achieved mean absolute errors (MAE) of 2.9-5.0 years. Essential differences between these three algorithms were (A) their training datasets, (B) preprocessing pipelines, and (C) deep learning architectures.

#### 2.3.1. Pyment

The Pyment algorithm (Leonardsen et al., 2022) was trained on 34,285 healthy individuals (age range 3 – 95 years) from 21 non-overlapping and publicly available neuroimaging datasets. T1-weighted data were acquired using a range of 1.5T and 3T scanners across different sites, using scanners from General Electric Healthcare, Siemens and Philips Healthcare. All MRI scans underwent QC procedures before inclusion, consisting of checks for preprocessing failures, as well as visual inspection. Preprocessing in Pyment consisted of skull-stripping with FreeSurfer 5.3 auto-recon pipeline (Segonne et al., 2004), reorienting to the standard FSL orientation, and linear registration to MNI152 with *flirt* using linear interpolation. Finally, the images were intensity normalized. The algorithm is based on a Simple Fully Convolutional Network (SFCN) with 6 convolutional blocks. The reported mean absolute error (MAE) for this algorithm was 5.0 years, and the reported R was 0.961. Code of used commit 00acac5 (v2.0.0) is available at: https://github.com/estenhl/pyment-public.

#### 2.3.2. MIDI

The MIDI algorithm (Wood et al., 2022) was trained on 2,387 clinical MRI scans, with an age range of 18 – 94 years, from two large hospitals, using a GE Signa 3T Discovery MR750 scanner with a variety of clinical scan protocols as used in routine NHS practice (Wood et al., 2022). All scans were reported radiologically normal for age without brain pathology, and no scans were excluded based on image quality. The original study did not report the number of patients and repeated visits (Wood et al., 2022). Preprocessing included skull-stripping using HD-BET (Isensee et al., 2019), followed by affine registration to MNI-space with ANTs, and intensity normalization. The algorithm is based on the DenseNet121 architecture (Wood et al., 2022). The reported MAE for this algorithm was 3.4 years and the reported R was 0.96. The code of used commit 028f4c6 is available at: https://github.com/MIDIconsortium/BrainAge.

#### 2.3.3. MCCQR

The MCCQR algorithm (Hahn et al., 2022) was trained on 10,691 healthy individuals (age range 20 – 75 years) from the German National Cohort (GNC) dataset. MRI scans were acquired across five sites equipped with dedicated identical MRI scanners (Skyra, Siemens Healthineers), using harmonized protocols across sites. Only scans of sufficient quality were included (Bamberg et al., 2015). Preprocessing for MCCQR was done with the CAT12 toolbox with default parameters. Images were bias corrected, segmented using tissue classification, normalized to MNI-space using DARTEL normalization, smoothed with an isotropic 8 mm Gaussian Kernel, and resampled to a 3-mm isotropic resolution. The algorithm architecture is based on a multi-channel 3D convolutional neural network combined with quantile regression, enabling both point estimates and uncertainty quantification of brain age. The reported MAE for this algorithm was 2.9 years, and R was not reported. Used code of commit 4a00072 and algorithm details are available at: https://github.com/wwu-mmll/uncertainty-brain-age.

### 2.4. Statistical analysis

All statistical analyses were conducted in R version 4.4.1.

#### 2.4.1. Deviations from artifact-free brain age predictions

The effects of artifacts and their severity on predicted brain age across all prediction algorithms were investigated with linear mixed-effect models. All models were fitted using the *lme4* package, with participant as a random intercept to account for artifact severities as repeated measures. We considered artifact severity as an unordered factor, enabling comparisons between artifact-free images (level 0) and each severity level separately (levels 1-10). Partial η² was calculated for the linear mixed-effect models, as it captures the proportion of variance explained by each effect or interaction. Partial η² was interpreted using conventional bins: very small (<0.02), small (0.02–0.13), medium (0.13–0.26), and large (>0.26) (Cohen, 2016).

Firstly, we investigated whether the effects of artifact severity on predicted brain age varied across algorithms and artifact types by modeling a three-way-interaction between brain age prediction algorithm (Pyment, MIDI, MCCQR), artifact type (motion, ghosting, blurring, noise) and severity level (0-10) on predicted brain age (outcome variable). Secondly, we conducted linear mixed effect models separately for each prediction algorithm, to test the interaction between artifact type and severity level.

Thirdly, post-hoc pairwise comparisons between severities (severity 1 vs 0, severity 2 vs 0, etc) were performed using the *emmeans* package, applying Benjamini-Hochberg FDR-corrections to account for multiple comparisons. Statistical significance was defined as two-tailed p < 0.05. Cohen’s *d* was computed for the post-hoc pairwise comparisons, where it provides a measure of how strongly each severity level deviates from the artifact-free condition. For both negative and positive values, Cohen’s *d* was interpreted using the following bins: negligible (< 0.20), small (0.20-0.50), medium (0.50-0.80), large (0.80-1.30), very large (> 1.30) (Coe, 2012).

#### 2.4.2. Prediction performance

Algorithm prediction performance was defined as the correspondence of predicted brain age to chronological age across all artifact types and severity levels. The following performance metrics were calculated using the *SimplyAgree* package: Pearson’s correlation coefficient (R), coefficient of determination (R^2^), MAE, and root mean squared error (RMSE). To facilitate comparisons of prediction performance across algorithms, both R and MAE were expressed as percentage change relative to artifact-free images (severity level 0).

Prediction performance analyses were additionally stratified by age to assess whether artifact-related effects were driven by specific age ranges (Zhang et al., 2023). Participants were grouped by their chronological age in three equally sized categories: younger adults (18–42 years), middle-aged adults (43–61 years), and older adults (62–79 years).

#### 2.4.3. Prediction stability

Algorithm prediction stability was assessed by intraclass correlation coefficients (ICC, two-way random-effects, absolute agreement) using the *irr* package, quantifying the consistency of brain age estimates within individuals across the eleven severity levels. ICC values were interpreted according to conventional thresholds: poor (<0.50), moderate (0.50-0.75), good (0.75-0.90), and excellent (>0.90) (Koo & Li, 2016).

In addition, we calculated the within-subject coefficient of variation (wsCV), providing a measure of relative variability in predicted brain age across artifact severities, expressed as a percentage, with lower scores indicating greater stability of the algorithm’s predictions. Differences between artifact-free and degraded images were examined using Bland-Altman analyses, calculating the mean difference in brain age gap (bias) and the limits of agreement (LoA; ±1.96 standard deviations of differences). Additionally, we assessed the percentage of brain age predictions falling within a maximal allowable difference (MAD) of ±3.55 years, following Hanson et al. (2024). This cutoff was based on clinically relevant brain age acceleration identified in a mega-analysis of patients with severe mental illness (Constantinides et al., 2023).

#### 2.4.4. Age bias correction

To investigate the impact of age-related bias on brain age estimates, we repeated all analyses using a standard statistical age-bias correction procedure. For each prediction algorithm, a linear regression was fitted between predicted brain age from artifact-free images and chronological age. Corrected predictions were calculated following **Fout! Verwijzingsbron niet gevonden.** (Cole et al., 2018):

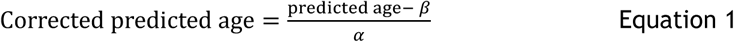

with α the slope, and β the intercept of the linear regression, respectively.

## 3. Results

### 3.1. Data inclusion across algorithms

A flowchart of participant inclusion across all algorithms and for each artifact and severity level is provided in **Suppl. Figure 1**. Not all degraded images (n = 2930) were successfully preprocessed for the Pyment and MCCQR algorithms. For the Pyment algorithm, preprocessing failures occurred for all artifact types, predominantly at higher severity levels. The largest number of unsuccessfully preprocessed images was observed for noise (n = 321), followed by blurring (n = 255), ghosting (n = 240), and motion (n = 194). For MCCQR, only higher severity ghosting gave unsuccessfully processed images (n = 11). All degraded images (n = 2930) were successfully pre-processed for the MIDI algorithm.

### 3.2. Deviations from artifact-free brain age predictions

In artifact-free scans, all three algorithms showed the following baseline performance (Pyment: R=0.92, R²=0.85, RMSE=8.6 years, MAE=7.1 years; MIDI: R=0.90, R²=0.80, RMSE=11.0 years, MAE=9.1 years; MCCQR: R=0.96, R²=0.92, RMSE=5.7 years, MAE=4.6 years**, Suppl. Table 2**).

The three-way linear mixed effects model revealed an interaction between artifact severity, brain age prediction algorithm, and artifact type (F(60, 37179) = 34.87, P < 0.0001; **Table 1**), indicating algorithm-specific sensitivity to different artifact types (**Figure 2**). The choice of brain age prediction algorithm accounted for the largest proportion of variance (η² = 0.39).

**Figure 2.**
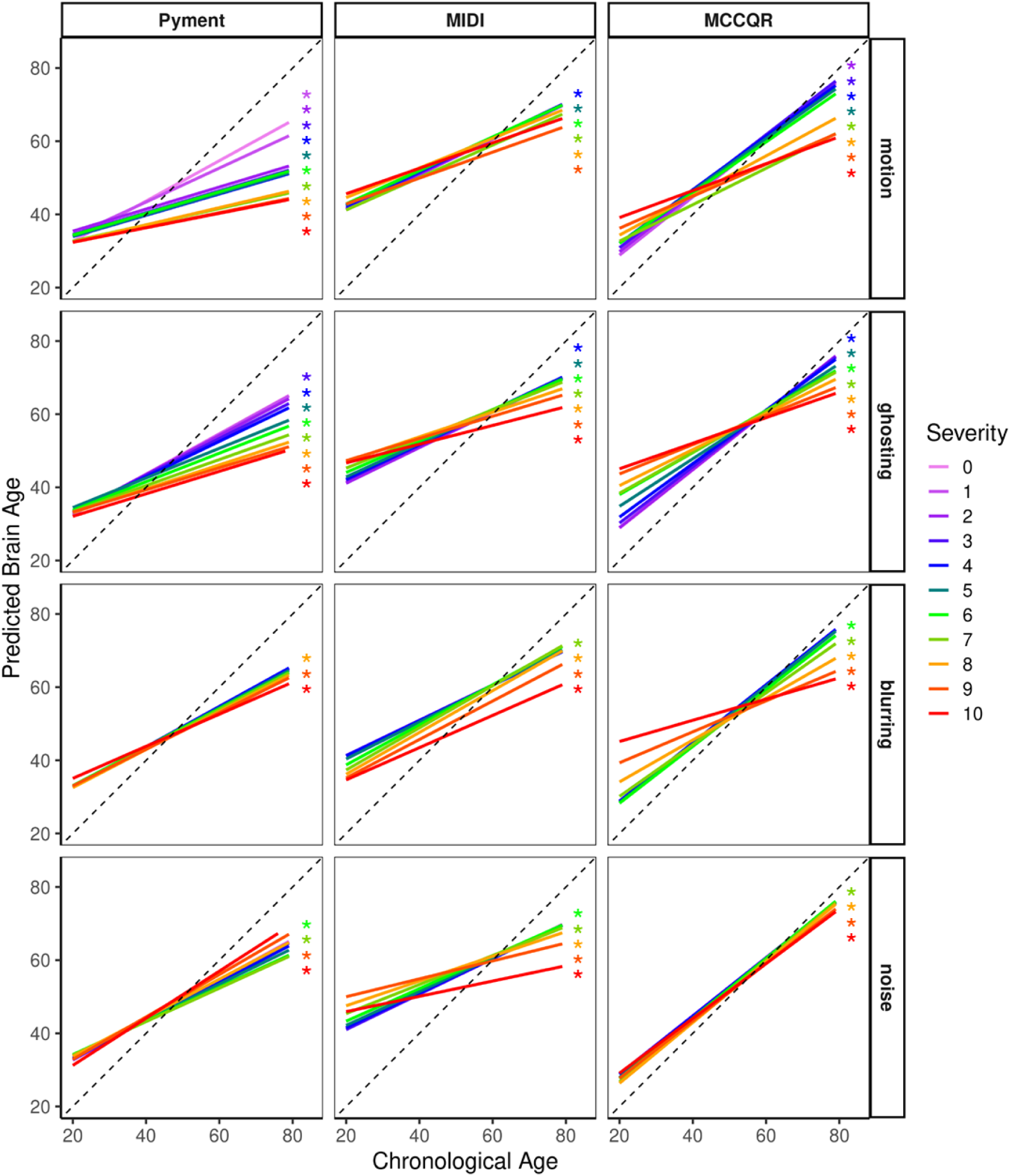
Predicted brain age across artifacts, algorithms, and severities. Predicted brain age as a function of chronological age, stratified by artifact type (rows), algorithm (columns), and artifact severity (colored lines). Each panel shows linear mixed-effects model fits for severity levels 0–10, with the dashed line indicating the identity line (predicted = chronological age). Asterisks mark severity levels whose predictions differed significantly from artifact-free images (p < 0.05), with the star color corresponding to the respective severity level.

**Table 1.** Main and interaction effects from the three-way-interaction linear mixed effects model.

| Linear mixed effects model | F statistic | p-value | $\eta^2$ |
| --- | --- | --- | --- |
| algorithm | ( 2,37180) = 11740.13 | <.0001 | 0.387 (large) |
| artifact | ( 3,37179) = 474.65 | <.0001 | 0.037 (small) |
| severity | (10,37179) = 126.92 | <.0001 | 0.033 (small) |
| algorithm*artifact | ( 6,37179) = 554.15 | <.0001 | 0.082 (small) |
| algorithm*severity | (20,37179) = 48.39 | <.0001 | 0.025 (small) |
| artifact*severity | (30,37179) = 31.57 | <.0001 | 0.025 (small) |
| algorithm*artifact*severity | (60,37179) = 34.87 | <.0001 | 0.053 (small) |

Linear mixed effects models for each brain age prediction algorithm separately, revealed significant effects of artifact type, severity and their interaction (all P < 0.0001, for all algorithms, **Table 2**). For Pyment, the effect of artifact type was large (η² = 0.374), whereas the effects of severity (η² = 0.172) and the severity × artifact interaction (η² = 0.186) were medium. In contrast, for MIDI and MCCQR, effect sizes for severity, artifact type, and their interaction were all small (η² = 0.03–0.123), suggesting less sensitivity to artifact type and severity than Pyment.

**Table 2.** Main and interaction effects from the post hoc two-way-interaction linear mixed effects models.

|  |  |  |  |
| --- | --- | --- | --- |
| A. Pyment |  |  |  |
| artifact | F( 3,11500) = 2294.09 | P<.0001 | $\eta^2$ = 0.374 (large) |
| severity | F(10,11498) = 239.32 | P<.0001 | $\eta^2$ = 0.172 (medium) |
| artifact*severity | F(30,11498) = 87.77 | P<.0001 | $\eta^2$ = 0.186 (medium) |
| B. MIDI |  |  |  |
| artifact | F( 3,12556) = 267.79 | P<.0001 | $\eta^2$ = 0.060 (small) |
| severity | F(10,12556) = 160.21 | P<.0001 | $\eta^2$ = 0.113 (small) |
| artifact*severity | F(30,12556) = 58.73 | P<.0001 | $\eta^2$ = 0.123 (small) |
| C. MCCQR |  |  |  |
| artifact | F( 3,12545) = 245.79 | P<.0001 | $\eta^2$ = 0.056 (small) |
| severity | F(10,12545) = 31.05 | P<.0001 | $\eta^2$ = 0.024 (small) |
| artifact*severity | F(30,12545) = 38.20 | P<.0001 | $\eta^2$ = 0.084 (small) |

Motion artifacts affected predicted brain age across all three algorithms, with the strongest effects observed for Pyment. For Pyment, deviations in predicted brain age from artifact-free images were significant from severity level 1 onward (P < 0.0001, **Suppl. Table 3A**) and Cohen’s d<-1.41 (very large) from severity level 2 onward (**Suppl. Table 3B**). In the MIDI algorithm, motion effects became significant from severity level 4 onward (**Suppl. Table 3C**), with some non-significant levels at severity 6 and 10, and effect sizes were negligible to small (d<0.29, **Suppl. Table 3D**). For MCCQR, significant deviations from artifact-free images were observed from severity level 2 onward, except for severity 6 (**Suppl. Table 3E**), with negligible to small effect sizes (d<0.36, **Suppl. Table 3F**).

Ghosting artifacts produced significant deviations from artifact-free images for Pyment from severity level 3 onward (**Suppl. Table 3A**) with large effect sizes from severity 7 onward (d<-1.50, **Suppl. Table 3B**). In the MIDI algorithm, ghosting effects became significant from severity level 4 onward (**Suppl. Table 3C**), with small or negligible effect sizes (d<0.43, **Suppl. Table 3D**). For MCCQR, significant deviations from artifact-free images were observed from severity level 4 onward (**Suppl. Table 3E**), with medium effect sizes starting from severity 6 (d>0.62, **Suppl. Table 3F**).

Blurring had a smaller impact across algorithms. For Pyment, significant effects were observed only from severity level 8 onward (**Suppl. Table 3A**), with small effect sizes (d<-0.01, **Suppl. Table 3B**). For the MIDI algorithm, blurring effects became significant from severity level 7 onward (**Suppl. Table 3C**), and with negligible effect sizes (d<0.01, **Suppl. Table 3D**). For MCCQR, significant deviations from artifact-free images were observed from severity level 6 onward (**Suppl. Table 3E**), with negligible effect sizes except at severity level 10 (d=0.20, **Suppl. Table 3F**).

Noise artifacts resulted in significant effects from severity level 6 onward for Pyment, except at severity level 8 (Suppl. Table 3**A**), with small effect sizes (d<0.38, **Suppl. Table 3B**). MIDI showed significant noise effects from severity level 6 onwards (**Suppl. Table 3C**), with small to medium effect sizes (d<0.58, **Suppl. Table 3D**). For MCCQR, noise effects appeared from severity level 7 onward (**Suppl. Table 3E**), with negligible effect sizes (d<0.01, **Suppl. Table 3F**).

Repeating the linear mixed-effect analyses using age-bias–corrected predictions revealed that the main effect of the brain age prediction algorithm was substantially reduced. In contrast, the effects of severity, artifact type, and their interactions were slightly increased (all F > 30, P < 0.0001, η² = 0.03–0.07, small, **Suppl. Table 4**). At the post-hoc level, Cohen’s *d* for pairwise comparisons between artifact severity levels (vs severity 0) decreased markedly for Pyment for all artifact types (**Suppl. Table 5**), whereas MIDI and MCCQR post-hoc effects remained essentially unchanged (**Suppl. Table 6 & 7**).

### 3.3. Prediction performance

The effects of artifact type and its severity on brain age prediction performance are summarized in **Figure 3** (percentage change relative to artifact-free images) and **Suppl. Table 2** (raw values).

**Figure 3.**
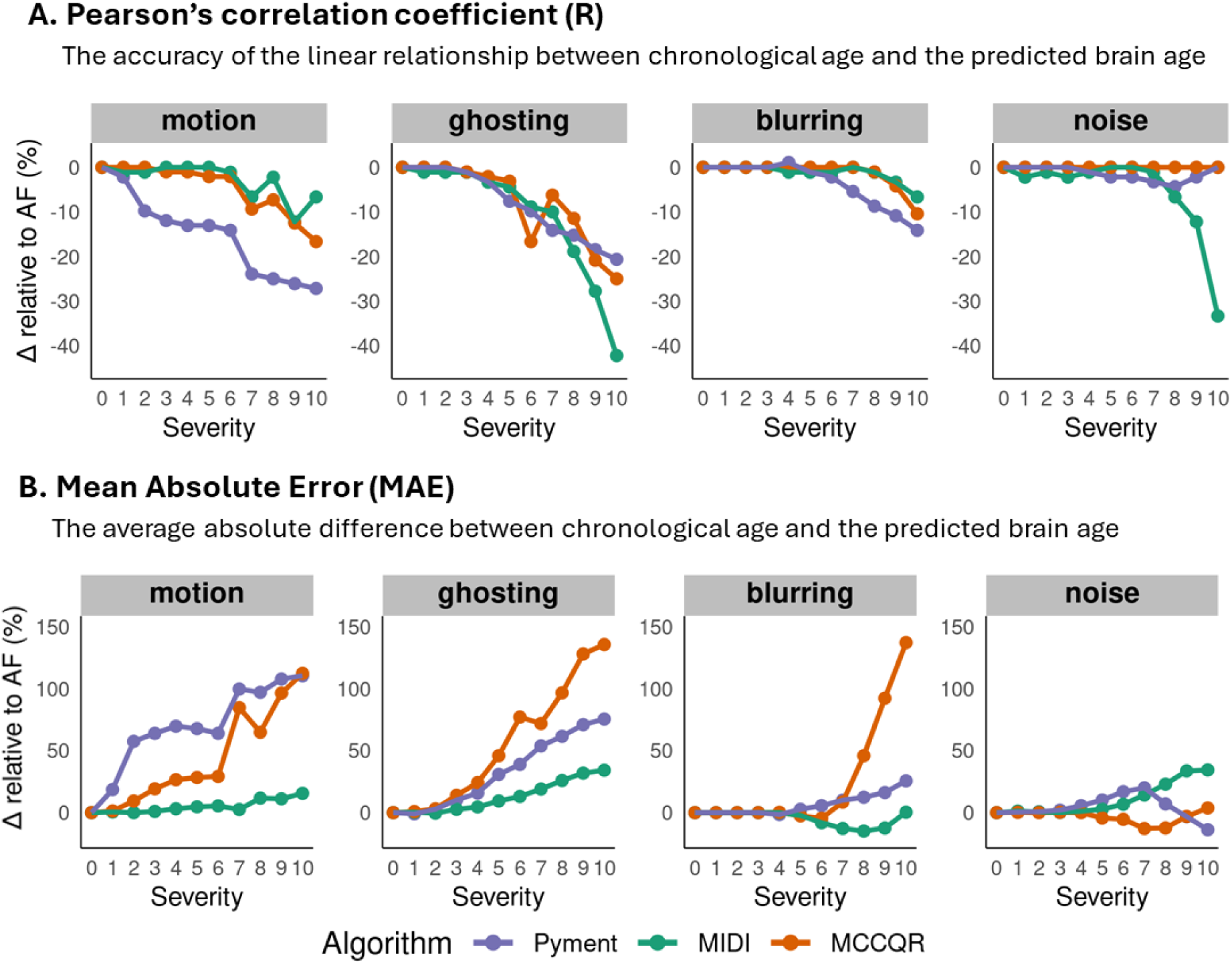
Performance metrics across artifact types and severity levels. Δ relative to AF (%) represents the percentage change in performance metrics (R and MAE) relative to the artifact-free condition (AF = severity level 0). The figure shows how prediction performance metrics change as artifact severity increases for each algorithm (Pyment, MIDI, MCCQR).

Motion artifacts led to progressive deterioration in prediction performance. For Pyment, correlations decreased strongly, with R decreasing with 27% at maximum severity. This was accompanied by an increase of 110% at maximum severity. For MIDI, we observed a decrease in R of 12%, and an MAE increase of 16% over the full range of severities. For MCCQR, we found a decrease in R of 17%, but a MAE increase of 112% at maximum severity.

Ghosting artifacts exerted the strongest influence on prediction performance. Pyment’s R declined with 21%, and MAE increased with 75% at maximum severity. For MIDI, we observed a decrease in R of 42%, and MAE increases of 34% over the full range of severities. For MCCQR, we found a decrease in R of 25%, but large MAE increases (135%) at maximum severity.

Blurring artifacts had minimal impact at low severity levels, but became progressively disruptive at higher severity levels. For Pyment, we found a decrease in R of 14%, and MAE increases of 26% at maximum severity. MIDI remained largely stable across all metrics, indicating robustness to blur. For MCCQR, we observed a decrease in R of 10%, accompanied by large MAE increases (137%) at severity level 10 Noise artifacts produced algorithm-specific patterns. Pyment was minimally affected, with a maximal decrease in R of 4%, and maximal MAE increase of 20%. For MIDI, we observed larger prediction performance declines, with an R decrease of 33% and MAE increase of 35% at maximum severity. MCCQR remained largely stable, with negligible changes in correlations and minor fluctuations in errors.

Age-dependent patterns in brain age prediction performance varied across algorithms and artifact types (**Figure 4**). For Pyment, older adults exhibited larger R decreases than young and middle-aged groups for motion, ghosting, and noise, whereas middle-aged adults showed the greatest performance degradation for blurring. Across all artifacts, Pyment demonstrated the largest increase in MAE in middle-aged groups. For MIDI, decreases in R were comparable across age groups, with similarly consistent MAE increases except at higher severities (8–10), where older adults exhibited markedly larger increases across all artifacts. MCCQR showed equivalent R drops across groups, but MAE increased most prominently in older adults for motion, blurring, and noise, whereas young adults displayed the greatest MAE elevations under ghosting.

**Figure 4.**
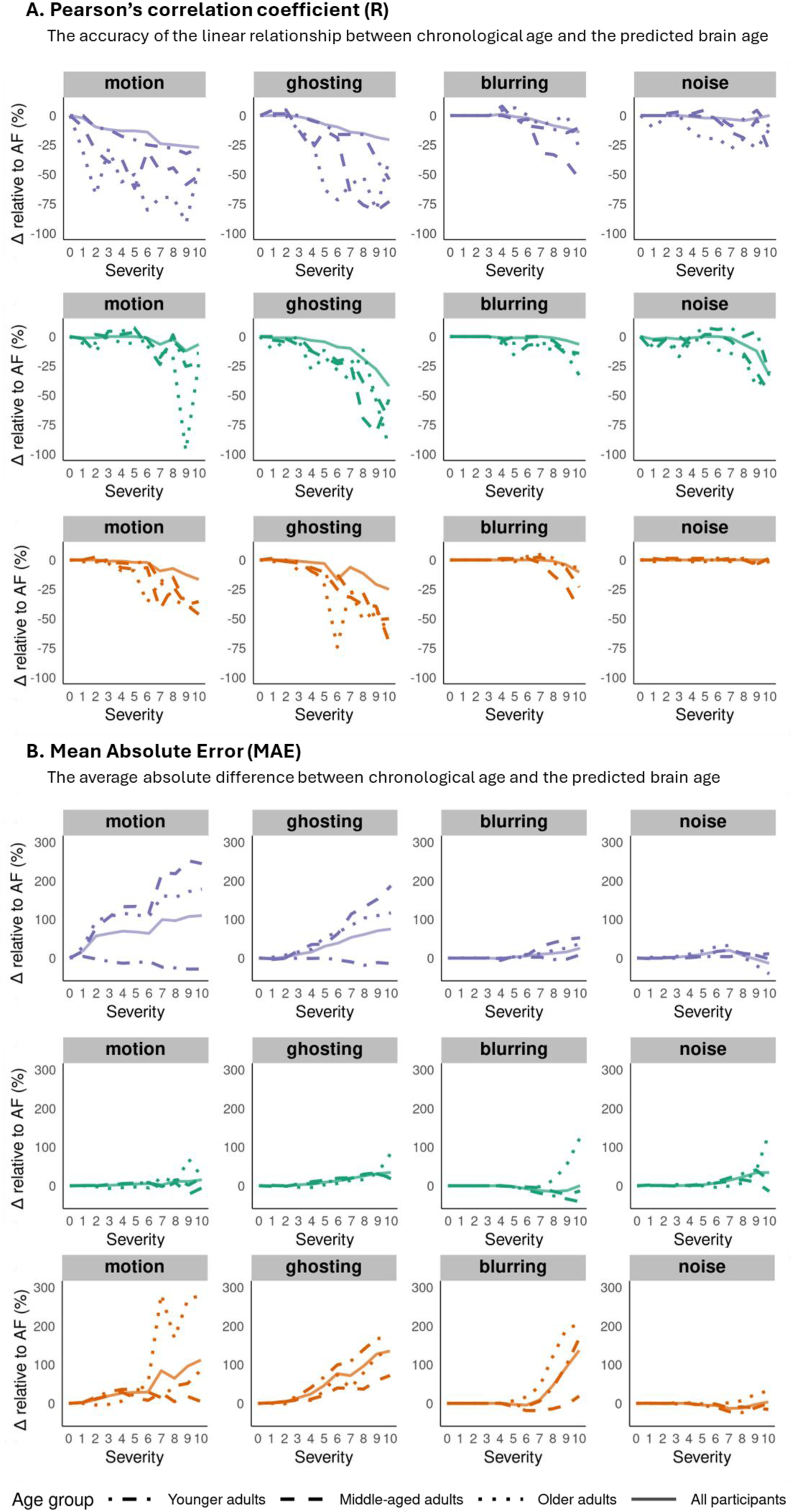
Performance metrics across artifact types and severity levels per age-group. Δ relative to AF (%) represents the percentage change in performance metrics (R and MAE) relative to the artifact-free condition (AF = severity level 0). The figure shows how prediction performance metrics change as artifact severity increases for each algorithm (Pyment, MIDI, MCCQR) across age-groups.

### 3.4. Prediction stability

The effects of artifact severity on brain age prediction stability are summarized in **Figure 5** (wsCV and ICC) and **Suppl. Figure 2** (Bland-Altman plots).

**Figure 5.**
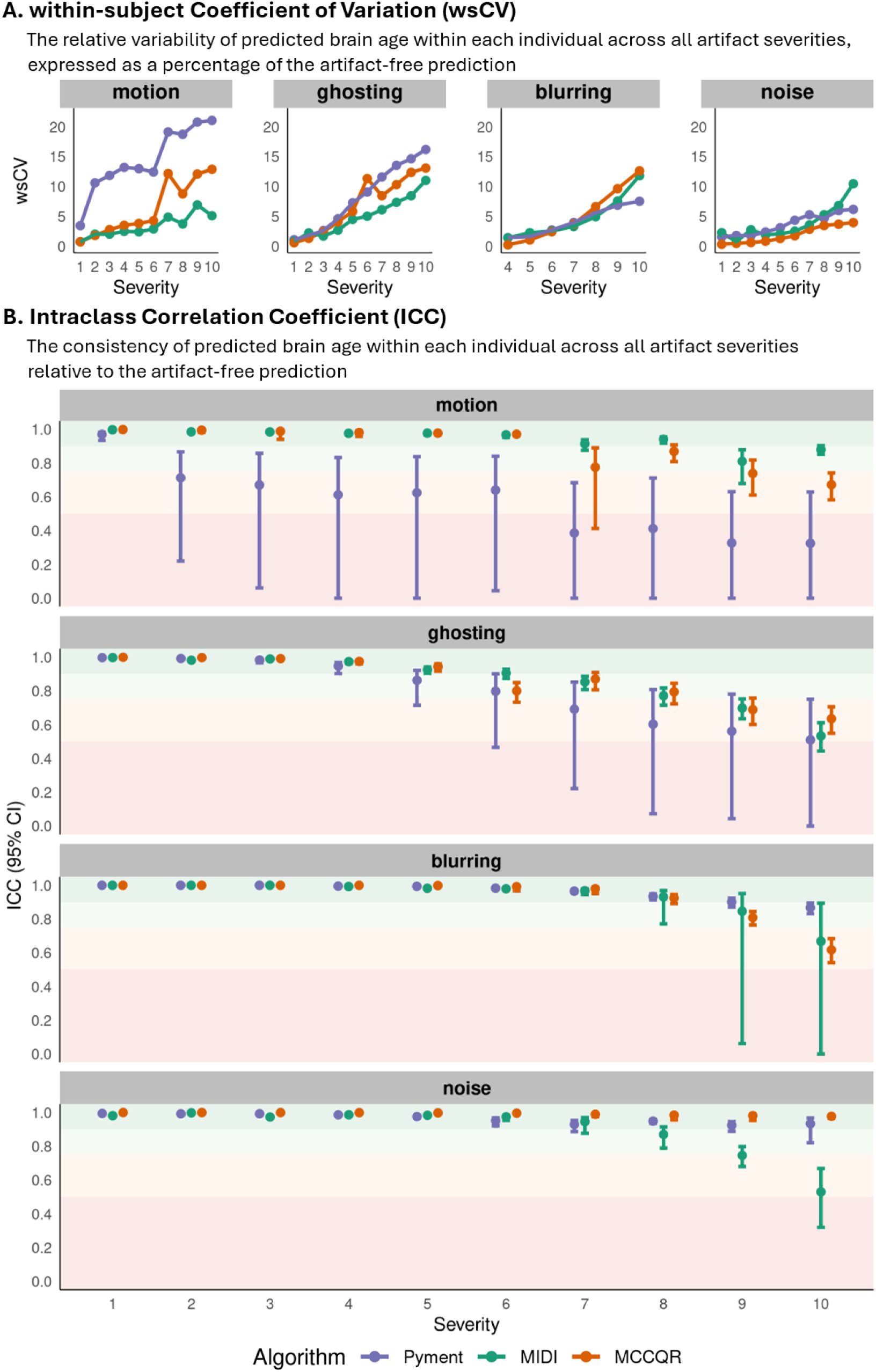
Impact of artifact severity on prediction stability. (A) Within-subject coefficient of variation (wsCV) across severity levels for each artifact type and algorithm (Pyment, MIDI, MCCQR). (B) Intraclass correlation coefficient (ICC) across severity levels, with shaded horizontal bands representing interpretive categories (Koo & Li, 2016): poor (<0.50, red), moderate (0.50–0.75, orange), good (0.75–0.90, light green), and excellent (>0.90, darker green). Points indicate estimates at each severity level, and vertical bars show 95% confidence intervals.

Motion artifacts progressively reduced prediction stability. For Pyment, ICC values declined from excellent to poor, with wsCV rising from 3.4% to 21.1%. Bland-Altman analysis showed increasing bias, and widening limits of agreement. The proportion of predictions within the MAD threshold dropped sharply, from 91% to 17%, reflecting substantial absolute errors. MIDI remained highly stable in terms of prediction stability, with ICC consistently good or excellent and wsCV below 7%. Bland-Altman analysis showed small bias (≤2 years) and >53% within MAD. MCCQR demonstrated excellent ICC at low severity levels, and declined to moderate at the highest two severities, with wsCV increasing from 0.7% to 12.9%. Biases and LoA widened moderately, and the proportion within MAD decreased from 100% to 30%.

Ghosting artifacts similarly impacted predicted stability. Pyment exhibited excellent ICC at low severity, decreasing to moderate at severity 10, with wsCV rising from 1.1% to 16.2%. Bias increased, LoA expanded, and percentage within MAD fell from 100% to 25%, indicating progressively degraded agreement. MIDI started with excellent ICC, and dropped to moderate with increasing severity, and wsCV increased from 0.8% to 11%. Bland-Altman analysis showed biases <.51 year, and the proportion within MAD decreasing from >99% to 39%. MCCQR showed excellent ICC initially, declining to moderate at highest severity; wsCV increased from 0.6% to 13%, and the proportion within MAD decreased from 100% to 31%.

Blurring artifacts minimally affected performance stability at low severity. Pyment retained excellent ICC and low wsCV (<7%) up to severity level 9. At severity level 10, ICC declined to 0.87 (good), wsCV increased to 7.5%, bias remained moderate (0.46 years), and the proportion within MAD decreased to 54%. MIDI followed similar trends, with ICC staying excellent and low wsCV until severity level 8. At the highest severity levels, ICC declined to 0.67, wsCV increased up to 11.8%, and MAD compliance dropped to 10%. MCCQR retained excellent ICC at mild blur, but moderate ICC (0.62) and wsCV 12.6% at highest severity indicated increasing variability, with absolute errors reflected in broader LoA and reduced MAD compliance (21%).

Noise artifacts generally had minimal impact on Pyment’s BA prediction, with ICC >0.92 across severities, wsCV <6.2%, and biases ≤2.6 years; MAD compliance remained >55%. MIDI exhibited more sensitivity at extreme noise, ICC decreased to 0.53 and wsCV increased to 10.45% at severity 10, with bias 3.6 years and only 36% of predictions within MAD. MCCQR was largely robust to noise, maintaining excellent ICC and low wsCV (<4%) across severities, with bias ≤1.24 years and MAD compliance >77%, indicating preserved stability despite increasing noise.

Repeating the performance analyses using age-bias–corrected predictions yielded only minor changes. For MIDI, MAE was slightly reduced at lower artifact severities, whereas Pyment showed small increases in MAE at higher severities (**Suppl. Table 8**). Importantly, the relative performance patterns across algorithms and artifact types were preserved, and reliability (ICC) remained in the same qualitative bins (**Suppl. Figure 3**). These results indicate that age-bias correction affected absolute metric values modestly but did not alter the overall sensitivity of predicted brain age to artifact type or severity.

## 4. Discussion

### 4.1. Summary of findings

This study yielded three main findings. First, differences in algorithms (pre-trained on clinical vs research datasets, preprocessing pipelines, algorithm architecture), led to differences in resilience to low-to-moderate artifact severities. Second, motion and ghosting artifacts induced substantial deviations from artifact-free brain age predictions, and the strongest declines in prediction performance and prediction stability. Motion and ghosting already showed significant effects on brain age prediction at low-to-moderate artifact severities (1-4), below the PondrAI QC threshold that would appear visually abnormal. Third, age bias and age-related effects shaped apparent artifact sensitivity, with age-bias correction strongly attenuating Pyment’s artifact effects and age-stratified analyses revealing stronger performance degradation in older adults for several artifacts and algorithms.

### 4.2. Artifact-specific effects

Motion and ghosting produced the strongest decline in brain-age prediction, the earliest statistical deviations from artifact-free brain-age predictions, as well as the strongest declines in prediction performance and prediction stability compared to blurring and noise artifacts. For Pyment, motion and ghosting consistently degraded both prediction performance (correlations dropped, MAE more than doubled) and prediction stability (ICC declined from excellent to moderate/poor, within-subject CV rose >10–20%), indicating predictions became both systematically biased and highly unstable across degraded scans. For MIDI and MCCQR, the performance prediction decreased broadly along the same lines as Pyment, but especially for motion the performance stability metrics showed less impaired prediction stability. This implies that these algorithms maintain more consistent predictions despite structured spatial distortions from motion and ghosting, while Pyment predictions fragment more severely under the same conditions. Both motion and ghosting artifacts create structured, spatially coherent distortions that disrupt the anatomical integrity of the image. Ghosting creates repeated signal patterns along the phase-encoding direction, and motion combines blurring, ghosting, and spurious signal changes that alter both spatial and intensity information (Atkinson et al., 1999; Vakli et al., 2024; Zaitsev et al., 2015). Such distortions can mimic biologically meaningful morphological variations, potentially misleading deep learning based algorithms trained to detect age-related anatomical changes (Cole & Franke, 2017; Tanveer et al., 2023). This may explain why motion and ghosting produced the largest performance degradations and greatest instability in brain age estimates. Such effects should be further investigated in patient cohorts, as differences between acute local and diffuse neurodegenerative pathology in spatial distributions can affect performance degradations differently.

In contrast, blurring and noise impacted brain age performance and prediction stability only at moderate to high severities, and effect sizes remained small for most algorithms. Blurring illustrated a partial decoupling between prediction performance and stability, particularly for MIDI and MCCQR. For MIDI, prediction performance changes were modest even at strong blur, whereas ICC dropped to moderate levels and wsCV and MAD violations increased, indicating that mean performance metrics understated the degree of prediction variability introduced by blur. MCCQR showed a similar pattern: prediction performance decreased only slightly under severe blurring, yet prediction stability metrics showed increasing instability, implying that relative ordering of brain age was largely preserved while absolute predictions became noisier and less reliable. For noise, Pyment and MCCQR no major declines in prediction performance and prediction stability were observed, suggesting that these algorithms treated the simulated noise largely as benign variability. By contrast, MIDI showed a pronounced vulnerability to high noise: at the highest severities, both prediction performance and prediction stability decreased, indicating that noise simultaneously impaired both ranking accuracy and stability for this algorithm. Blurring and noise locally perturb voxel intensities while preserving image structure. Blurring is a low-pass spatial filter that reduces image sharpness and fine texture, while Gaussian noise introduces random intensity fluctuations (Khan et al., 2019). The relative robustness to low-to-moderate blurring and noise suggests that current brain-age algorithms depend more strongly on global structural features than on fine-grained, high-frequency details, in line with prior work showing that classical machine-learning algorithms achieve competitive brain-age performance (Da Costa et al., 2020).

All artifacts in this study were simulated using TorchIO (Perez-Garcia et al., 2021), allowing controlled yet stochastic simulations of artifact severity. For motion, both the magnitude and number of transformations increased with severity, while their directions and temporal order were randomly sampled; ghosting intensity was scaled proportionally but with random phase and axis selection. As a result, higher severity levels simulated not only stronger but also more spatially heterogeneous distortions. This increased stochasticity at higher artifact severities likely contributed to the increased variability in brain age performance observed at high artifact levels. Although these artifacts already appear realistic, recent work argued that commonly used magnitude-based simulations paradigms, such as those implemented in TorchIO, do not fully replicate the spatial complexity and anatomical disruptions associated with real head motion in structural MRI (Olsson et al., 2024). As a result, the effects of motion artifacts on brain-age predictions reported here are likely conservative with respect to real-world settings. Future work employing anatomically more realistic simulation pipelines may yield greater fidelity in estimating the impact of motion and other artifacts on neuroimaging-derived brain measures.

### 4.3. Differences between brain age algorithms

The prediction algorithms different substantially in their resilience to artifacts, which may reflect differences between their training data, pre-processing pipelines, and deep learning architecture. While we cannot fully disentangle the contribution of these components from the present data, we can appreciate some patterns. Whereas Pyment and MCCQR were trained on healthy individuals included in research studies, MIDI was trained on patient scans that were radiologically normal for age without brain pathology. All three algorithms resampled the images to standard space, although only Pyment and MIDI performed skull-stripping. Pyment’s preprocessing pipeline, calibrated largely on aggregated research-quality T1-weighted datasets, showed heightened vulnerability to image artifact-induced failures compared to MIDI, and to some extent MCCQR. This agrees with prior machine-learning based work showing that algorithms trained on research-centric samples, characterized by stringent diagnostic exclusions, standardized acquisition protocols, and thorough quality control, perform well on idealized external research cohorts but struggle to generalize to the variability encountered in clinical practice (Baecker et al., 2021; Jónsson et al., 2019; Xiong et al., 2023). In contrast, clinical datasets encompass a broader spectrum of age ranges, scanners, health statuses, and artifact prevalence, fostering an pipeline resilience reflected here in MIDI’s comparatively stable performance when challenged with diverse artifact types. Finally, the heavy reliance on UK Biobank data in Pyment’s training set (roughly 75%), which predominantly originate from a single scanner type and single image protocol, may have contributed to its reduced generalizability and increased vulnerability to simulated artifacts.

Architecturally, the Pyment algorithm relies on the SFCN (Peng et al., 2021), a comparatively streamlined architecture with relatively few parameters and strong regularization. DenseNet architectures, such as the one underlying MIDI, utilize dense skip connections that propagate low-level features to deeper layers, enabling more redundant representation learning and potentially better buffering against localized image corruption (Zhang et al., 2021). This architectural trait may additionally explain MIDI’s relative stability in the presence of mild blurring and noise. While the literature reports mixed evidence regarding the superiority of more complex architectures in brain age prediction, with some studies indicating that increasing complexity does not uniformly guarantee improved robustness or generalizability (Hendrycks & Dietterich, 2019; Keremis et al., 2025), our results suggests that the increased architecture complexity of DenseNet outperforms SFCN in unseen image distortions.

Complementing these architectural features, the MCCQR algorithm employs composite quantile regression with Monte Carlo dropout to explicitly quantify both aleatory and epistemic uncertainty, which provides an additional mechanism to buffer against prediction bias introduced by artifacts (Gal & Ghahramani, 2016). In our results, MCCQR showed the strongest baseline accuracy, relatively small statistical deviations for most artifact types, and excellent to moderate prediction stability across a wide range of severities, but it also displayed marked MAE inflation under the most severe for motion, ghosting, and blurring levels. This pattern suggests that MCCQR preserved rank ordering of age reasonably well but exhibited substantial shifts in absolute brain-age estimates under extreme structural distortions, leading to large absolute errors despite moderate correlations. These findings indicate that MCCQR’s uncertainty-aware framework may not fully prevent bias when the input images depart strongly from its training domain, even though its relative ranking performance remains robust.

### 4.4. Preprocessing failures

Preprocessing failures were a major source of data loss and revealed algorithm-specific sensitivities to artifact type and severity. Pyment showed the highest preprocessing failure rate, particularly for blurring and noise, possibly reflecting the dependency of the FreeSurfer-based skull-stripping and registration steps on image contrast and sharp anatomical boundaries (Waters et al., 2019). In contrast, MIDI’s preprocessing pipeline completed successfully for all degraded images, and MCCQR’s pipeline only failed for a small subset of high-severity ghosting images. Although the linear mixed-effects models can accommodate unequal group sizes, the reduced sample for Pyment likely contributed to less stable parameter estimates and may have led to an overestimation of Pyment’s performance at extreme artifact severities, because the most severely degraded images were selectively removed. An alternative solution could have been the simulation of artifacts in the standard space, after the preprocessing of non-degraded images. This could have avoided algorithm-and artifact-specific preprocessing failures. However, this would bypass the way in which preprocessing steps are affected by artifacts present in native images, thereby reducing validity.

Furthermore, each brain-age algorithm was coupled to its own preprocessing pipeline. These pipelines may interact differently with specific artifact types, making it complicated to attribute performance declines solely to the underlying network of the prediction algorithm itself as opposed to upstream preprocessing instabilities. Since no additional quality control was performed on preprocessing outputs beyond successful pipeline completion, a subset of prediction errors may therefore reflect preprocessing variability rather than inherent limitations of the brain age algorithms.

### 4.5. Age bias correction and age-stratified artifact sensitivity

Repeating all analyses with age-bias-corrected predictions showed that age bias primarily inflated apparent artifact sensitivity in the Pyment algorithm, whereas there was no age bias effect for MIDI and MCCQR. After correction, Pyment’s post-hoc effects were remarkedly reduced across all artifact types, indicating that a portion of Pyment’s previously observed sensitivity was explained by age-dependent prediction bias rather than true degradation effects.

Age-stratified analyses of performance degradation further revealed that the sensitivity to artifacts differed by algorithm and artifact type rather than following a single global age effect. This aligned with established training biases in brain age algorithms, as many brain age algorithms are trained on datasets with uneven age distributions, often underrepresenting the oldest age ranges, which may amplify sensitivity to domain shift when older brains are further degraded by artifacts (Bashyam et al., 2020). In addition, aging-related reductions in tissue contrast and cortical thinning resulting in partial volume effects may decrease the structural image information content for brain age, potentially relatively increasing the artifact effects. From a clinical perspective, these results are relevant because brain age prediction is increasingly applied to aging and neurodegenerative cohorts. The age-dependent vulnerability underscores the need for artifact-robust and age-balanced training datasets.

### 4.6. Towards clinically robust brain-age estimations

Our findings underscore that the fact that current brain age algorithms are predominantly validated on high-quality research datasets, limits their translational readiness, and emphasizes the need for quality-agnostic or quality-aware algorithms that can operate robustly across variable clinical MRI acquisition environments (Gaser et al., 2024). To start, artifact-free/non-degraded images (level 0) already show large differences in prediction performance that affect clinical application. In Alzheimer’s Disease (AD), where brain age is commonly applied, mild cognitive impairment (BAG<4 years) and dementia stages (BAG>5 years) exhibit age gaps smaller than the algorithm performance of all included algorithms (Liu et al., 2025). In these clinical applications, BAGs and BAG variability are more pronounced and will be substantially affected by the artifacts like those simulated in the present work, highlighting the need for additional evaluations of prediction performance in clinical-grade data and the effects of artifacts on brain age predictions.

A critical focus of future work should be to determine the artifact severity threshold at which brain age predictions lose their clinical utility. This threshold is reached when artifact-induced prediction errors exceed the typical deviations observed between healthy controls and clinical populations. Gaps below ∼3.5 years are typically considered negligible, whereas larger deviations signal potential pathology. Thus, when simulated artifacts reduce MAD compliance, they introduce variability or biases that either mask clinical signals or produce spurious accelerations mimicking disease, rendering the brain age prediction uninterpretable for individual-level diagnosis. Furthermore, many diseases start locally, while general aging has a more uniform pattern (Bethlehem et al., 2022) and could therefore be differently affected by the artefacts examined in this study. For example, in (late-onset) AD, hippocampal volume shows early atrophy (Meysami et al., 2021) and rotational motion may have less effect on such a central region within the head compared to outer regions where motion has more pronounced effects. Pyment’s rapid compliance decline below 30% under motion by severity 2 indicates this research-optimized algorithm fails at moderate degradations commonly encountered in clinical settings, where even visually neglectable artifacts push predictions beyond clinically interpretable bounds and risk false disease. MIDI maintains >70% compliance up to higher severities across all simulated artifacts, indicating clinical-grade training data value for routine robustness, though its sharp blurring drop-off suggests architectural limits to global smoothing tolerance. MCCQR balances resilience across artifacts, sustaining >75% for noise and blurring while matching Pyment’s motion/ghosting vulnerability. Interpretability thus hinges on both artifact type and algorithm choice. Furthermore, algorithm explainability is currently not considered, but it is of high importance to understand the applicability of deep-learning-based models in the clinic, and could provide a better window into how artifacts affect brain age predictions. Such assessments could help to understand which models would be better suited to explore altered ageing trajectories in specific diseases and determine if artifacts would affect brain age estimates.

### 4.7. Limitations

One limitation of this study is that our artifact simulations did not capture the full complexity of real-world MRI variability. For example, hardware-related artifacts and more complex composite artifacts were not simulated and artefacts were generated slice-wise rather than in 3D. Consequently, the reported effects likely represent conservative estimates of the impact of real-world artifacts. This motivates two complementary directions: extending simulation frameworks to more faithfully reproduce empirically observed clinical artifacts, and pairing such simulations with multi-center clinical datasets that include detailed artifact annotations, enabling direct comparison between simulated and real-world failures.

The ABRIM dataset provided research-grade T1-weighted scans from a neurologically healthy sample spanning a broad adult age range, offering a controlled setting in which the isolated impact of simulated artifacts on brain-age predictions could be examined with minimal confounding from pre-existing pathology or pre-existing image artifacts. At the same time, this level of standardization and health homogeneity limits the generalizability as it is not yet known how simulated artifacts would interact with variable scanner hardware, protocols, and comorbidities.

Additionally, our evaluation focused primarily on a cross-sectional dataset obtained using a single scanner; longitudinal assessments are needed to determine how artifact sensitivity affects the stability and consistency of brain-age estimates over time.

### 4.8. Conclusion

Our findings demonstrate that simulated MRI artifacts differentially affect brain age prediction depending on artifact type, severity, and algorithm, with motion and ghosting producing the largest disruptions. The vulnerability of different algorithms highlights that both preprocessing strategies and underlying architectures influence robustness, emphasizing that artifact sensitivity is a key consideration when interpreting brain-age as a biomarker. Our study encourages the design of artifact-aware calibration strategies. Importantly, training on diverse clinical datasets and implementing robust preprocessing pipelines may improve resilience, underscoring practical strategies to enhance reliability and facilitate translation to real-world clinical and multi-site imaging settings.

Future work should incorporate real-world artifact data to capture the full spectrum of clinically encountered distortions. Finally, investigating the downstream clinical impact of artifact-induced errors by measuring brain age sensitivity and specificity against confirmed pathology in patient cohorts, will be crucial to establish practical performance thresholds and inform clinical deployment.

## Data Availability

All original data is available at https://data.ru.nl/collections/di/dccn/DSC_3015046.06_419. All data produced in the present study are avilable upon reasonable request to the authors https://data.ru.nl/collections/di/dccn/DSC_3015046.06_419

## Supplemental methods

The TorchIO artifact simulation parameter values and range used for artifact generation were based on visual quality ratings, and were such that they correspond to the full PondrAI QC range. To emphasize the lower quality scores, we implemented a power function transformation:

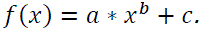

Here, *x* represents the severity level, whereas *f*(*x*) represents the corresponding artifact parameter value. The function was parameterized using three reference points for each artifact type, which were mapped onto a coordinate system to structure the severity scale. The starting point was defined as (0,0), where *x* = 0 corresponds to the artifact-free images. The severity scale was then defined in 11 steps, progressing from 0 (artifact-free) to *x* = 10 (maximum artifact severity). To structure the scale, we used three reference points: the artifact-free images (*x* = 0, serving as the true minimum), moderate artifact severity (*x* = 7, corresponding to PondrAI QC score 3, good image quality), and maximum artifact severity (*x* = 10 corresponding to PondrAI QC score 6, severely degraded images). A schematic representation of these reference points, in correspondence to the PondrAI QC scores, is displayed in **Figure 1A**.

To establish these reference points empirically (*x* = 7, and *x* = 10), we used N = 40,000 augmented MRI scans from our previous work as a starting point (Hendriks et al., 2025), in which we simulated four artifact types (motion, ghosting, blurring, and noise) at ten severity levels based on (Loizillon et al., 2024). From this dataset, we randomly selected n = 100 MRI scans, ensuring balanced representation across artifact types and severities. Subsequently, two independent experts (JH, MGJ) manually rated these scans using PondrAI QC guidelines. Since subtle MRI degradations may remain undetectable to human raters, we expected that some augmented images would receive a perfect rating despite containing minor artifacts. The resulting ratings were then used to assess whether the predefined artifact severity levels adequately reflected the PondrAI QC score range, from score 1 (perfect) to score 6 (terribly degraded image quality).

In cases where the initial severity levels did not sufficiently capture this range, the parameter range was adjusted to improve representation, introducing either more or less severe artifacts where necessary. To facilitate consensus on the final reference points for each artifact type (*x* = 7, and *x* = 10), we sampled 100 severity levels across n = 3 subjects, from “perfect” to severely degraded images. A consensus meeting was then held to determine the reference points that would be embedded in the power function transformation. More specifically, for each artifact type, a system of equations was defined based on the empirically defined reference points, and the parameters *a*, *b*, and *c* were solved numerically. These parameters were then used to compute the artifact parameter value across the ten severity levels.

## Supplemental Tables

**Suppl. Table 1.** Torchio parameter values specifying the magnitude of motion, ghosting, blurring, and noise artifacts at each severity level (0–10) for MRI simulations.

| Severity level | motion | ghosting | blurring | noise |
| --- | --- | --- | --- | --- |
| 0 | 0.000 | 0.000 | 0.000 | 0.000 |
| 1 | 0.068 | 0.030 | 0.005 | 0.001 |
| 2 | 0.263 | 0.095 | 0.032 | 0.005 |
| 3 | 0.578 | 0.186 | 0.091 | 0.017 |
| 4 | 1.011 | 0.298 | 0.190 | 0.040 |
| 5 | 1.560 | 0.431 | 0.337 | 0.078 |
| 6 | 2.223 | 0.582 | 0.538 | 0.133 |
| 7 | 3.000 | 0.750 | 0.800 | 0.210 |
| 8 | 3.889 | 0.935 | 1.127 | 0.311 |
| 9 | 4.889 | 1.135 | 1.526 | 0.440 |
| 10 | 6.000 | 1.350 | 2.000 | 0.600 |

**Suppl. Table 2.** Prediction performance metrics for brain age algorithms (Pyment, MIDI, MCCQR) across simulated MRI artifact types and severity levels. Reported metrics include Pearson’s correlation coefficient (R), coefficient of determination (R²), root mean squared error (RMSE), and mean absolute error (MAE).

|  |  | Pyment |  |  |  | MIDI |  |  |  | MCCQR |  |  |  |
| --- | --- | --- | --- | --- | --- | --- | --- | --- | --- | --- | --- | --- | --- |
| artifact | severity | R | $R^2$ | RMSE | MAE | R | $R^2$ | RMSE | MAE | R | $R^2$ | RMSE | MAE |
| artifact-free | 0 | 0.92 | 0.85 | 8.57 | 7.11 | 0.9 | 0.8 | 10.98 | 9.08 | 0.96 | 0.92 | 5.67 | 4.6 |
| motion | 1 | 0.90 | 0.80 | 10.09 | 8.43 | 0.89 | 0.80 | 11.05 | 9.13 | 0.96 | 0.92 | 5.73 | 4.66 |
| motion | 2 | 0.83 | 0.68 | 13.58 | 11.19 | 0.89 | 0.79 | 11.06 | 9.08 | 0.96 | 0.92 | 6.19 | 5.03 |
| motion | 3 | 0.81 | 0.66 | 14.06 | 11.65 | 0.90 | 0.82 | 11.15 | 9.17 | 0.95 | 0.91 | 6.75 | 5.48 |
| motion | 4 | 0.80 | 0.64 | 14.58 | 12.06 | 0.90 | 0.81 | 11.36 | 9.36 | 0.95 | 0.90 | 7.10 | 5.82 |
| motion | 5 | 0.80 | 0.63 | 14.39 | 11.92 | 0.90 | 0.81 | 11.50 | 9.50 | 0.94 | 0.89 | 7.22 | 5.90 |
| motion | 6 | 0.79 | 0.62 | 14.14 | 11.65 | 0.89 | 0.79 | 11.65 | 9.57 | 0.94 | 0.89 | 7.14 | 5.94 |
| motion | 7 | 0.70 | 0.48 | 17.24 | 14.19 | 0.84 | 0.71 | 11.34 | 9.31 | 0.87 | 0.75 | 10.14 | 8.49 |
| motion | 8 | 0.69 | 0.48 | 17.08 | 14.00 | 0.88 | 0.78 | 12.32 | 10.14 | 0.89 | 0.79 | 9.10 | 7.58 |
| motion | 9 | 0.68 | 0.46 | 18.00 | 14.76 | 0.79 | 0.62 | 12.28 | 10.09 | 0.84 | 0.70 | 10.88 | 9.03 |
| motion | 10 | 0.67 | 0.45 | 18.28 | 14.95 | 0.84 | 0.71 | 12.93 | 10.49 | 0.80 | 0.64 | 11.80 | 9.76 |
| ghosting | 1 | 0.92 | 0.85 | 8.49 | 7.04 | 0.89 | 0.80 | 11.06 | 9.11 | 0.96 | 0.92 | 5.70 | 4.64 |
| ghosting | 2 | 0.92 | 0.85 | 8.78 | 7.31 | 0.89 | 0.79 | 10.98 | 9.04 | 0.96 | 0.92 | 5.86 | 4.74 |
| ghosting | 3 | 0.91 | 0.83 | 9.34 | 7.83 | 0.89 | 0.79 | 11.33 | 9.32 | 0.95 | 0.91 | 6.43 | 5.24 |
| ghosting | 4 | 0.89 | 0.79 | 9.83 | 8.23 | 0.87 | 0.77 | 11.60 | 9.49 | 0.94 | 0.89 | 7.25 | 5.71 |
| ghosting | 5 | 0.85 | 0.73 | 11.39 | 9.30 | 0.86 | 0.74 | 11.95 | 9.93 | 0.93 | 0.86 | 8.36 | 6.71 |
| ghosting | 6 | 0.83 | 0.69 | 12.09 | 9.87 | 0.82 | 0.68 | 12.58 | 10.26 | 0.80 | 0.64 | 11.43 | 8.14 |
| ghosting | 7 | 0.79 | 0.62 | 13.24 | 10.93 | 0.81 | 0.65 | 13.04 | 10.80 | 0.90 | 0.80 | 10.03 | 7.90 |
| ghosting | 8 | 0.78 | 0.61 | 14.12 | 11.48 | 0.73 | 0.54 | 14.05 | 11.43 | 0.85 | 0.72 | 11.28 | 9.04 |
| ghosting | 9 | 0.75 | 0.57 | 14.77 | 12.15 | 0.65 | 0.42 | 14.55 | 11.97 | 0.76 | 0.58 | 12.89 | 10.48 |
| ghosting | 10 | 0.73 | 0.53 | 15.10 | 12.47 | 0.52 | 0.27 | 15.02 | 12.18 | 0.72 | 0.52 | 13.37 | 10.83 |
| blurring | 1 | 0.92 | 0.85 | 8.57 | 7.11 | 0.90 | 0.80 | 10.98 | 9.08 | 0.96 | 0.92 | 5.67 | 4.60 |
| blurring | 2 | 0.92 | 0.85 | 8.57 | 7.11 | 0.90 | 0.80 | 10.98 | 9.08 | 0.96 | 0.92 | 5.67 | 4.60 |
| blurring | 3 | 0.92 | 0.85 | 8.57 | 7.11 | 0.90 | 0.80 | 10.98 | 9.08 | 0.96 | 0.92 | 5.67 | 4.60 |
| blurring | 4 | 0.93 | 0.86 | 8.45 | 7.00 | 0.89 | 0.79 | 10.99 | 9.08 | 0.96 | 0.92 | 5.68 | 4.61 |
| blurring | 5 | 0.91 | 0.84 | 8.77 | 7.30 | 0.89 | 0.78 | 10.77 | 8.87 | 0.96 | 0.93 | 5.51 | 4.48 |
| blurring | 6 | 0.90 | 0.81 | 9.05 | 7.51 | 0.89 | 0.80 | 10.15 | 8.34 | 0.96 | 0.93 | 5.41 | 4.39 |
| blurring | 7 | 0.87 | 0.75 | 9.42 | 7.82 | 0.90 | 0.81 | 9.58 | 7.92 | 0.96 | 0.92 | 6.04 | 4.99 |
| blurring | 8 | 0.84 | 0.71 | 9.74 | 7.99 | 0.89 | 0.79 | 9.32 | 7.73 | 0.95 | 0.89 | 7.93 | 6.71 |
| blurring | 9 | 0.82 | 0.67 | 10.22 | 8.26 | 0.87 | 0.76 | 9.58 | 7.95 | 0.92 | 0.84 | 10.35 | 8.84 |
| blurring | 10 | 0.79 | 0.63 | 10.82 | 8.93 | 0.84 | 0.70 | 11.08 | 9.11 | 0.86 | 0.74 | 12.82 | 10.90 |
| noise | 1 | 0.92 | 0.84 | 8.69 | 7.14 | 0.88 | 0.77 | 11.11 | 9.19 | 0.96 | 0.92 | 5.66 | 4.61 |
| noise | 2 | 0.92 | 0.84 | 8.62 | 7.16 | 0.89 | 0.80 | 11.07 | 9.13 | 0.96 | 0.92 | 5.68 | 4.60 |
| noise | 3 | 0.92 | 0.84 | 8.68 | 7.25 | 0.88 | 0.77 | 11.02 | 9.09 | 0.96 | 0.92 | 5.67 | 4.61 |
| noise | 4 | 0.91 | 0.83 | 9.05 | 7.51 | 0.89 | 0.79 | 11.13 | 9.17 | 0.96 | 0.92 | 5.66 | 4.60 |
| noise | 5 | 0.90 | 0.82 | 9.48 | 7.83 | 0.90 | 0.81 | 11.29 | 9.33 | 0.96 | 0.93 | 5.45 | 4.40 |
| noise | 6 | 0.90 | 0.80 | 9.99 | 8.31 | 0.90 | 0.81 | 11.78 | 9.69 | 0.96 | 0.92 | 5.35 | 4.35 |
| noise | 7 | 0.89 | 0.79 | 10.28 | 8.53 | 0.89 | 0.79 | 12.66 | 10.37 | 0.96 | 0.93 | 5.04 | 4.01 |
| noise | 8 | 0.88 | 0.78 | 9.29 | 7.61 | 0.84 | 0.71 | 13.68 | 11.16 | 0.96 | 0.93 | 4.99 | 4.03 |
| noise | 9 | 0.90 | 0.82 | 8.56 | 6.88 | 0.79 | 0.62 | 14.77 | 12.13 | 0.96 | 0.92 | 5.51 | 4.46 |
| noise | 10 | 0.92 | 0.84 | 7.61 | 6.13 | 0.60 | 0.36 | 14.35 | 12.20 | 0.96 | 0.91 | 5.90 | 4.77 |

**Suppl. Table 3.** Post-hoc comparisons of artifact severity effects on predicted brain age across algorithms. P-values (A, C, E) and corresponding Cohen’s d effect sizes (B, D, F) are shown for pairwise contrasts between each severity level (1–10) and the artifact-free condition (0) for Pyment, MIDI, and MCCQR. Significant values indicate levels of severity that produce deviations from artifact-free predictions.

|  | Severity 1 | Severity 2 | Severity 3 | Severity 4 | Severity 5 | Severity 6 | Severity 7 | Severity 8 | Severity 9 | Severity 10 |
| --- | --- | --- | --- | --- | --- | --- | --- | --- | --- | --- |
| motion | P<.0001 | P<.0001 | P<.0001 | P<.0001 | P<.0001 | P<.0001 | P<.0001 | P<.0001 | P<.0001 | P<.0001 |
| ghosting | P=0.77 | P=0.35 | P=0.0052 | P<.0001 | P<.0001 | P<.0001 | P<.0001 | P<.0001 | P<.0001 | P<.0001 |
| blurring | P=0.93 | P=0.93 | P=0.93 | P=0.93 | P=0.93 | P=0.36 | P=0.08 | P=0.0007 | P=0.0001 | P=0.0016 |
| noise | P=0.92 | P=0.92 | P=0.74 | P=0.33 | P=0.07 | P=0.0001 | P<.0001 | P=0.33 | P<.0001 | P<.0001 |

B. Post hoc analysis (effect sizes, Cohen's *d*) - Pymment
|  | Severity 1 | Severity 2 | Severity 3 | Severity 4 | Severity 5 | Severity 6 | Severity 7 | Severity 8 | Severity 9 | Severity 10 |
| --- | --- | --- | --- | --- | --- | --- | --- | --- | --- | --- |
| motion | d = -0.35 | d = -1.41 | d = -1.70 | d = -1.95 | d = -1.90 | d = -1.77 | d = -2.89 | d = -2.86 | d = -3.16 | d = -3.19 |
| ghosting | d = -0.02 | d = -0.08 | d = -0.24 | d = -0.42 | d = -0.75 | d = -1.10 | d = -1.50 | d = -1.84 | d = -2.00 | d = -2.24 |
| blurring | d = -0.01 | d = -0.01 | d = -0.01 | d = 0.01 | d = -0.05 | d = -0.11 | d = -0.18 | d = -0.33 | d = -0.38 | d = -0.30 |
| noise | d = -0.01 | d = -0.01 | d = -0.05 | d = -0.10 | d = -0.18 | d = -0.35 | d = -0.41 | d = 0.11 | d = 0.38 | d = 0.54 |

C. Post hoc analysis (p values, *emmeans* contrasts) - MIDI
|  | Severity 1 | Severity 2 | Severity 3 | Severity 4 | Severity 5 | Severity 6 | Severity 7 | Severity 8 | Severity 9 | Severity 10 |
| --- | --- | --- | --- | --- | --- | --- | --- | --- | --- | --- |
| motion | P=0.96 | P=0.96 | P=0.15 | P=0.033 | P=0.016 | P=0.0035 | P<.0001 | P=0.0014 | P<.0001 | P=0.47 |
| ghosting | P=0.99 | P=0.99 | P=0.13 | P=0.015 | P=0.0038 | P<.0001 | P<.0001 | P<.0001 | P=0.05 | P<.0001 |
| blurring | P>.99 | P>.99 | P>.99 | P>.99 | P=0.67 | P=0.08 | P<.0001 | P<.0001 | P<.0001 | P<.0001 |
| noise | P=0.70 | P=0.96 | P=0.54 | P=0.96 | P=0.47 | P=0.0009 | P<.0001 | P<.0001 | P<.0001 | P<.0001 |

D. Post hoc analysis (effect sizes, Cohen's *d*) - MIDI
|  | Severity 1 | Severity 2 | Severity 3 | Severity 4 | Severity 5 | Severity 6 | Severity 7 | Severity 8 | Severity 9 | Severity 10 |
| --- | --- | --- | --- | --- | --- | --- | --- | --- | --- | --- |
| motion | d = 0.01 | d = <0.01 | d = 0.13 | d = 0.19 | d = 0.22 | d = 0.26 | d = <0.01 | d = 0.29 | d = <0.01 | d = 0.07 |
| ghosting | d = <0.01 | d = <0.01 | d = 0.14 | d = 0.22 | d = 0.26 | d = 0.37 | d = 0.43 | d = 0.42 | d = 0.17 | d = <0.01 |
| blurring | d = <0.01 | d = <0.01 | d = <0.01 | d = <0.01 | d = <0.01 | d = <0.01 | d = <0.01 | d = <0.01 | d = <0.01 | d = <0.01 |
| noise | d = <0.01 | d = 0.01 | d = <0.01 | d = <0.01 | d = 0.09 | d = 0.29 | d = 0.48 | d = 0.58 | d = 0.45 | d = <0.01 |

E. Post hoc analysis (p values, *emmeans* contrasts) - MCCQR
|  | Severity 1 | Severity 2 | Severity 3 | Severity 4 | Severity 5 | Severity 6 | Severity 7 | Severity 8 | Severity 9 | Severity 10 |
| --- | --- | --- | --- | --- | --- | --- | --- | --- | --- | --- |
| motion | P=0.75 | P=0.014 | P<.0001 | P<.0001 | P=0.011 | P=0.67 | P<.0001 | P<.0001 | P<.0001 | P<.0001 |
| ghosting | P=0.93 | P=0.51 | P=0.06 | P=0.0005 | P<.0001 | P<.0001 | P<.0001 | P<.0001 | P<.0001 | P<.0001 |
| blurring | P>.99 | P>.99 | P>.99 | P>.99 | P=0.34 | P=0.0017 | P<.0001 | P<.0001 | P=0.0062 | P=0.031 |
| noise | P=0.97 | P=0.97 | P=0.97 | P=0.97 | P=0.65 | P=0.40 | P=0.0020 | P<.0001 | P<.0001 | P=0.0020 |

**Suppl. Table 4.** Main and interaction effects from the three-way-interaction linear mixed effects model after age bias correction.

| Linear mixed effects model | F statistic | p-value | $\eta^2$ |
| --- | --- | --- | --- |
| algorithm | F(2,37180)=1516.86 | P<.0001 | $\eta^2=0.075$ (small) |
| artifact | F(3,37179)=592.76 | P<.0001 | $\eta^2=0.046$ (small) |
| severity | F(10,37179)=174.19 | P<.0001 | $\eta^2=0.045$ (small) |
| algorithm*artifact | F(6,37179)=750.61 | P<.0001 | $\eta^2=0.108$ (small) |
| algorithm*severity | F(20,37179)=73.95 | P<.0001 | $\eta^2=0.038$ (small) |
| artifact*severity | F(30,37179)=39.00 | P<.0001 | $\eta^2=0.031$ (small) |
| algorithm*artifact*severity | F(60,37179)=45.47 | P<.0001 | $\eta^2=0.068$ (small) |

**Suppl. Table 5.**
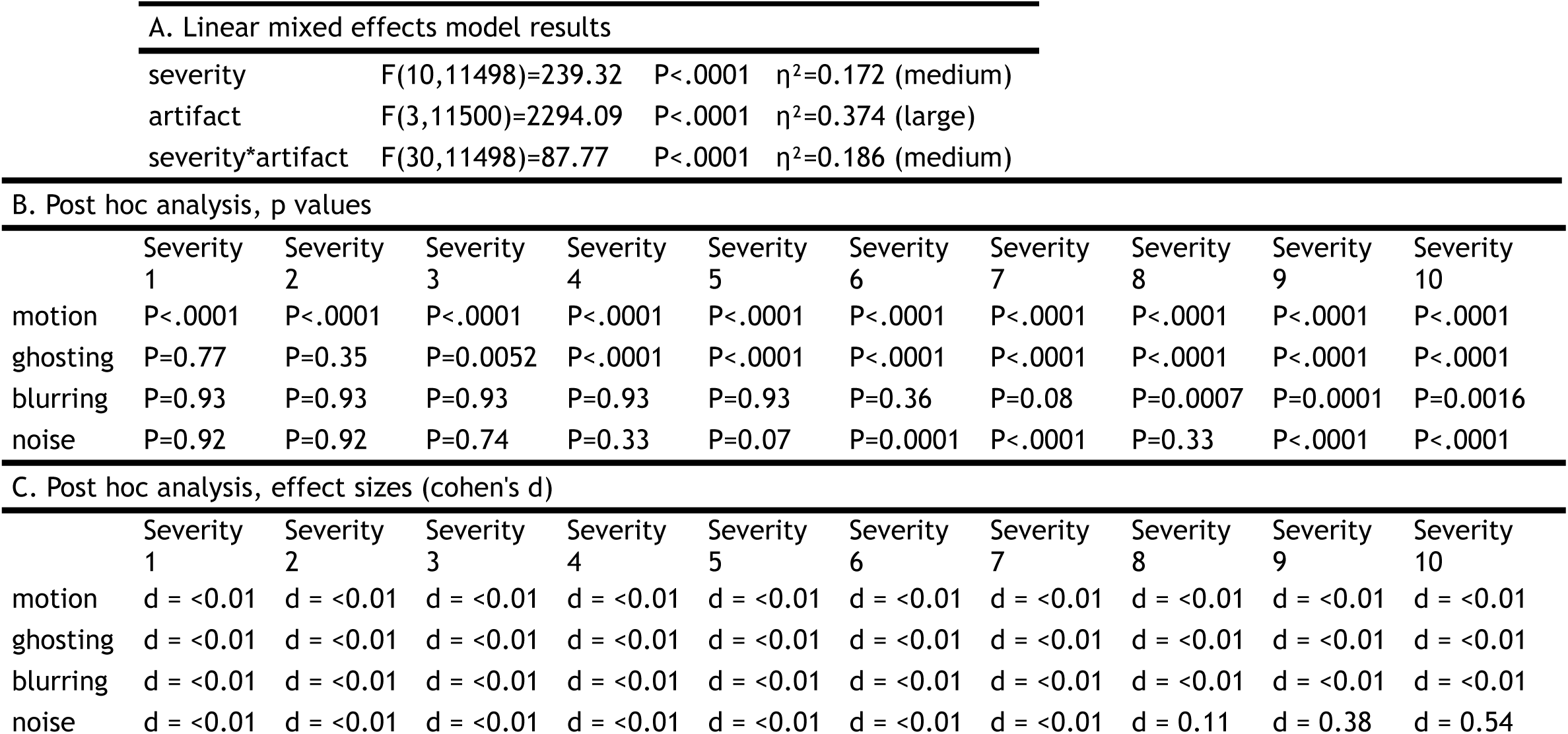
Post hoc analysis of severity effects by artifact after age bias correction (two-way model, Pyment)

A. Linear mixed effects model results
|  |  |  |  |
| --- | --- | --- | --- |
| severity | F(10,11498)=239.32 | P<.0001 | $\eta^2=0.172$ (medium) |
| artifact | F(3,11500)=2294.09 | P<.0001 | $\eta^2=0.374$ (large) |
| severity*artifact | F(30,11498)=87.77 | P<.0001 | $\eta^2=0.186$ (medium) |

B. Post hoc analysis, p values
|  | Severity<br>1 | Severity<br>2 | Severity<br>3 | Severity<br>4 | Severity<br>5 | Severity<br>6 | Severity<br>7 | Severity<br>8 | Severity<br>9 | Severity<br>10 |
| --- | --- | --- | --- | --- | --- | --- | --- | --- | --- | --- |
| motion | P<.0001 | P<.0001 | P<.0001 | P<.0001 | P<.0001 | P<.0001 | P<.0001 | P<.0001 | P<.0001 | P<.0001 |
| ghosting | P=0.77 | P=0.35 | P=0.0052 | P<.0001 | P<.0001 | P<.0001 | P<.0001 | P<.0001 | P<.0001 | P<.0001 |
| blurring | P=0.93 | P=0.93 | P=0.93 | P=0.93 | P=0.93 | P=0.36 | P=0.08 | P=0.0007 | P=0.0001 | P=0.0016 |
| noise | P=0.92 | P=0.92 | P=0.74 | P=0.33 | P=0.07 | P=0.0001 | P<.0001 | P=0.33 | P<.0001 | P<.0001 |

C. Post hoc analysis, effect sizes (cohen's d)
|  | Severity<br>1 | Severity<br>2 | Severity<br>3 | Severity<br>4 | Severity<br>5 | Severity<br>6 | Severity<br>7 | Severity<br>8 | Severity<br>9 | Severity<br>10 |
| --- | --- | --- | --- | --- | --- | --- | --- | --- | --- | --- |
| motion | d = <0.01 | d = <0.01 | d = <0.01 | d = <0.01 | d = <0.01 | d = <0.01 | d = <0.01 | d = <0.01 | d = <0.01 | d = <0.01 |
| ghosting | d = <0.01 | d = <0.01 | d = <0.01 | d = <0.01 | d = <0.01 | d = <0.01 | d = <0.01 | d = <0.01 | d = <0.01 | d = <0.01 |
| blurring | d = <0.01 | d = <0.01 | d = <0.01 | d = <0.01 | d = <0.01 | d = <0.01 | d = <0.01 | d = <0.01 | d = <0.01 | d = <0.01 |
| noise | d = <0.01 | d = <0.01 | d = <0.01 | d = <0.01 | d = <0.01 | d = <0.01 | d = <0.01 | d = 0.11 | d = 0.38 | d = 0.54 |

**Suppl. Table 6.** Post hoc analysis of severity effects by artifact after age bias correction (two-way model, MIDI)

| A. Linear mixed effects model results |  |  |  |
| --- | --- | --- | --- |
| severity | F(10,12556)=160.21 | P<0.001 | $\eta^2=0.113$ (small) |
| artifact | F(3,12556)=267.79 | P<.0001 | $\eta^2=0.060$ (small) |
| severity*artifact | F(30,12556)=58.73 | P<.0001 | $\eta^2=0.123$ (small) |

|  | Severity<br>1 | Severity<br>2 | Severity<br>3 | Severity<br>4 | Severity<br>5 | Severity<br>6 | Severity<br>7 | Severity<br>8 | Severity<br>9 | Severity<br>10 |
| --- | --- | --- | --- | --- | --- | --- | --- | --- | --- | --- |
| motion | P=0.96 | P=0.96 | P=0.15 | **P=0.033** | **P=0.016** | **P=0.0035** | **P<.0001** | **P=0.0014** | **P<.0001** | P=0.47 |
| ghosting | P=0.99 | P=0.99 | P=0.13 | **P=0.015** | **P=0.0038** | **P<.0001** | **P<.0001** | **P<.0001** | **P=0.05** | **P<.0001** |
| blurring | P>.99 | P>.99 | P>.99 | P>.99 | P=0.67 | P=0.08 | **P<.0001** | **P<.0001** | **P<.0001** | **P<.0001** |
| noise | P=0.70 | P=0.96 | P=0.54 | P=0.96 | P=0.47 | **P=0.0009** | **P<.0001** | **P<.0001** | **P<.0001** | **P<.0001** |

**Suppl. Table 7.** Post hoc analysis of severity effects by artifact after age bias correction (two-way model, MCCQR)

| A. Linear mixed effects model results |  |  |  |
| --- | --- | --- | --- |
| severity | F(10,12545)=31.05 | P<.0001 | $\eta^2=0.024$ (small) |
| artifact | F(3,12545)=245.79 | P<.0001 | $\eta^2=0.056$ (small) |
| severity*artifact | F(30,12545)=38.20 | P<.0001 | $\eta^2=0.084$ (small) |

|  | Severity<br>1 | Severity<br>2 | Severity<br>3 | Severity<br>4 | Severity<br>5 | Severity<br>6 | Severity<br>7 | Severity<br>8 | Severity<br>9 | Severity<br>10 |
| --- | --- | --- | --- | --- | --- | --- | --- | --- | --- | --- |
| motion | P=0.75 | **P=0.014** | **P<.0001** | **P<.0001** | **P=0.011** | P=0.67 | **P<.0001** | **P<.0001** | **P<.0001** | **P<.0001** |
| ghosting | P=0.93 | P=0.51 | **P=0.06** | **P=0.0005** | **P<.0001** | **P<.0001** | **P<.0001** | **P<.0001** | **P<.0001** | **P<.0001** |
| blurring | P>.99 | P>.99 | P>.99 | P>.99 | P=0.34 | **P=0.0017** | **P<.0001** | **P<.0001** | **P=0.0062** | **P=0.031** |
| noise | P=0.97 | P=0.97 | P=0.97 | P=0.97 | P=0.65 | P=0.40 | **P=0.0020** | **P<.0001** | **P<.0001** | **P=0.0020** |

**Suppl. Table 8.** Prediction performance metrics after age-bias correction for brain age algorithms (Pyment, MIDI, MCCQR) across simulated MRI artifact types and severity levels. Reported metrics include Pearson’s correlation coefficient (R), coefficient of determination (R²), root mean squared error (RMSE), and mean absolute error (MAE).

|  |  | Pyment |  |  |  | MIDI |  |  |  | MCCQR |  |  |  |
| --- | --- | --- | --- | --- | --- | --- | --- | --- | --- | --- | --- | --- | --- |
| artifact | severity | R | $R^2$ | RMSE | MAE | R | $R^2$ | RMSE | MAE | R | $R^2$ | RMSE | MAE |
| artifact-free | 0 | 0.92 | 0.85 | 7.22 | 5.72 | 0.90 | 0.80 | 8.40 | 6.48 | 0.96 | 0.92 | 4.88 | 3.78 |
| motion | 1 | 0.90 | 0.80 | 7.91 | 6.24 | 0.89 | 0.80 | 8.47 | 6.56 | 0.96 | 0.92 | 4.93 | 3.80 |
| motion | 2 | 0.83 | 0.68 | 13.10 | 10.44 | 0.89 | 0.79 | 8.95 | 6.70 | 0.96 | 0.92 | 5.04 | 3.96 |
| motion | 3 | 0.81 | 0.66 | 14.47 | 11.78 | 0.90 | 0.82 | 8.09 | 6.42 | 0.95 | 0.91 | 5.44 | 4.32 |
| motion | 4 | 0.80 | 0.64 | 15.73 | 12.92 | 0.90 | 0.81 | 8.27 | 6.45 | 0.95 | 0.90 | 5.56 | 4.43 |
| motion | 5 | 0.80 | 0.63 | 15.49 | 12.74 | 0.90 | 0.81 | 8.21 | 6.44 | 0.94 | 0.89 | 5.76 | 4.65 |
| motion | 6 | 0.79 | 0.62 | 14.92 | 11.94 | 0.89 | 0.79 | 8.48 | 6.65 | 0.94 | 0.89 | 5.69 | 4.66 |
| motion | 7 | 0.70 | 0.48 | 21.34 | 18.15 | 0.84 | 0.71 | 10.49 | 7.96 | 0.87 | 0.75 | 11.03 | 9.00 |
| motion | 8 | 0.69 | 0.48 | 21.17 | 17.90 | 0.88 | 0.78 | 8.29 | 6.61 | 0.89 | 0.79 | 8.58 | 6.82 |
| motion | 9 | 0.68 | 0.46 | 22.83 | 19.48 | 0.79 | 0.62 | 11.84 | 9.19 | 0.84 | 0.70 | 10.87 | 8.86 |
| motion | 10 | 0.67 | 0.45 | 23.20 | 19.69 | 0.84 | 0.71 | 9.25 | 7.34 | 0.80 | 0.64 | 11.49 | 9.22 |
| ghosting | 1 | 0.92 | 0.85 | 6.94 | 5.52 | 0.89 | 0.80 | 8.57 | 6.56 | 0.96 | 0.92 | 4.92 | 3.80 |
| ghosting | 2 | 0.92 | 0.85 | 6.84 | 5.45 | 0.89 | 0.79 | 8.83 | 6.66 | 0.96 | 0.92 | 4.94 | 3.88 |
| ghosting | 3 | 0.91 | 0.83 | 7.40 | 5.77 | 0.89 | 0.79 | 8.79 | 6.80 | 0.95 | 0.91 | 5.24 | 4.18 |
| ghosting | 4 | 0.89 | 0.79 | 8.21 | 6.29 | 0.87 | 0.77 | 9.36 | 7.41 | 0.94 | 0.89 | 5.86 | 4.58 |
| ghosting | 5 | 0.85 | 0.73 | 9.97 | 7.77 | 0.86 | 0.74 | 9.83 | 7.41 | 0.93 | 0.86 | 6.70 | 5.33 |
| ghosting | 6 | 0.83 | 0.69 | 11.56 | 9.02 | 0.82 | 0.68 | 11.04 | 8.48 | 0.80 | 0.64 | 11.03 | 6.78 |
| ghosting | 7 | 0.79 | 0.62 | 13.80 | 10.94 | 0.81 | 0.65 | 11.15 | 8.62 | 0.90 | 0.80 | 8.36 | 6.47 |
| ghosting | 8 | 0.78 | 0.61 | 15.40 | 12.14 | 0.73 | 0.54 | 12.51 | 9.84 | 0.85 | 0.72 | 9.81 | 7.90 |
| ghosting | 9 | 0.75 | 0.57 | 16.60 | 13.46 | 0.65 | 0.42 | 14.08 | 11.01 | 0.76 | 0.58 | 11.73 | 9.48 |
| ghosting | 10 | 0.73 | 0.53 | 17.90 | 14.86 | 0.52 | 0.27 | 17.05 | 12.38 | 0.72 | 0.52 | 12.36 | 9.86 |
| blurring | 1 | 0.92 | 0.85 | 7.22 | 5.72 | 0.90 | 0.80 | 8.40 | 6.48 | 0.96 | 0.92 | 4.88 | 3.78 |
| blurring | 2 | 0.92 | 0.85 | 7.22 | 5.72 | 0.90 | 0.80 | 8.40 | 6.48 | 0.96 | 0.92 | 4.88 | 3.78 |
| blurring | 3 | 0.92 | 0.85 | 7.22 | 5.72 | 0.90 | 0.80 | 8.40 | 6.48 | 0.96 | 0.92 | 4.88 | 3.78 |
| blurring | 4 | 0.93 | 0.86 | 6.92 | 5.57 | 0.89 | 0.79 | 8.69 | 6.64 | 0.96 | 0.92 | 4.90 | 3.81 |
| blurring | 5 | 0.91 | 0.84 | 7.38 | 5.86 | 0.89 | 0.78 | 9.42 | 6.95 | 0.96 | 0.93 | 4.86 | 3.74 |
| blurring | 6 | 0.90 | 0.81 | 7.93 | 6.29 | 0.89 | 0.80 | 9.95 | 7.57 | 0.96 | 0.93 | 4.87 | 3.74 |
| blurring | 7 | 0.87 | 0.75 | 9.37 | 7.34 | 0.90 | 0.81 | 10.47 | 8.19 | 0.96 | 0.92 | 5.03 | 3.96 |
| blurring | 8 | 0.84 | 0.71 | 10.44 | 8.23 | 0.89 | 0.79 | 11.72 | 9.44 | 0.95 | 0.89 | 6.65 | 5.47 |
| blurring | 9 | 0.82 | 0.67 | 11.28 | 8.94 | 0.87 | 0.76 | 14.27 | 11.70 | 0.92 | 0.84 | 8.98 | 7.69 |
| blurring | 10 | 0.79 | 0.63 | 10.99 | 8.67 | 0.84 | 0.70 | 19.08 | 16.52 | 0.86 | 0.74 | 11.51 | 9.90 |
| noise | 1 | 0.92 | 0.84 | 7.34 | 5.71 | 0.88 | 0.77 | 9.15 | 6.81 | 0.96 | 0.92 | 4.87 | 3.77 |
| noise | 2 | 0.92 | 0.84 | 7.23 | 5.79 | 0.89 | 0.80 | 8.46 | 6.58 | 0.96 | 0.92 | 4.92 | 3.81 |
| noise | 3 | 0.92 | 0.84 | 7.08 | 5.60 | 0.88 | 0.77 | 9.25 | 6.87 | 0.96 | 0.92 | 4.91 | 3.81 |
| noise | 4 | 0.91 | 0.83 | 7.37 | 5.75 | 0.89 | 0.79 | 8.63 | 6.55 | 0.96 | 0.92 | 4.94 | 3.80 |
| noise | 5 | 0.90 | 0.82 | 7.41 | 5.75 | 0.90 | 0.81 | 8.02 | 6.31 | 0.96 | 0.93 | 4.93 | 3.82 |
| noise | 6 | 0.90 | 0.80 | 7.80 | 6.01 | 0.90 | 0.81 | 7.96 | 6.30 | 0.96 | 0.92 | 5.08 | 3.92 |
| noise | 7 | 0.89 | 0.79 | 8.23 | 6.47 | 0.89 | 0.79 | 8.38 | 6.72 | 0.96 | 0.93 | 5.36 | 4.05 |
| noise | 8 | 0.88 | 0.78 | 8.61 | 6.51 | 0.84 | 0.71 | 9.89 | 7.95 | 0.96 | 0.93 | 5.38 | 4.22 |
| noise | 9 | 0.90 | 0.82 | 8.78 | 6.83 | 0.79 | 0.62 | 11.13 | 8.82 | 0.96 | 0.92 | 5.40 | 4.28 |
| noise | 10 | 0.92 | 0.84 | 8.90 | 7.21 | 0.60 | 0.36 | 15.82 | 12.71 | 0.96 | 0.91 | 5.19 | 4.18 |

## Supplemental Figures

**Suppl. Figure 1.**
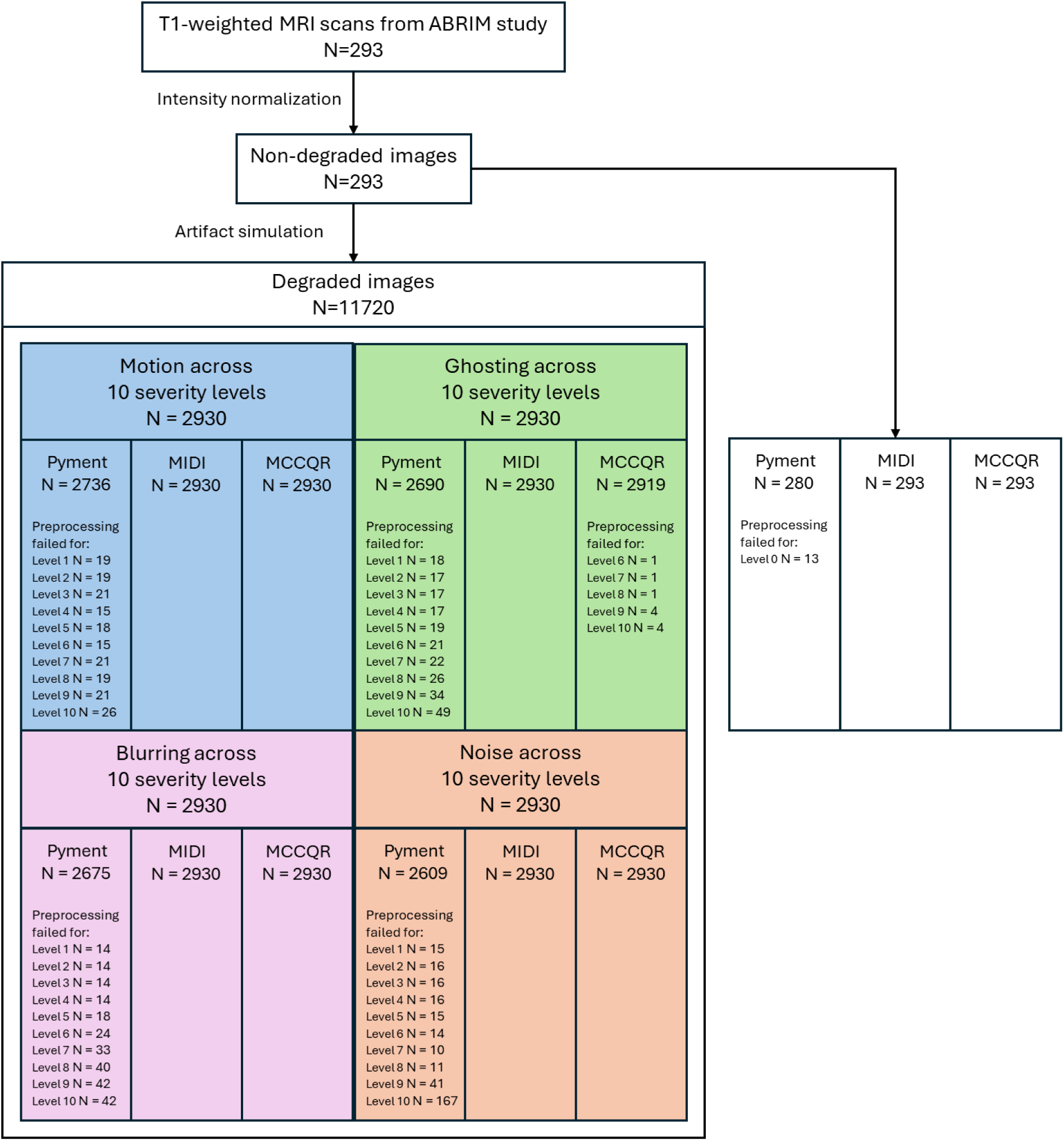
Flowchart of participant inclusion and preprocessing failures across artifact simulations and brain age algorithms. Flowchart showing the selection of T1-weighted MRI scans and the generation of degraded images for artifact simulation at 11 severity levels. The number of processed and unprocessed images is reported for each brain age prediction algorithm (Pyment, MIDI, MCCQR) and artifact type (motion, ghosting, blurring, noise).

**Suppl. Figure 2.**
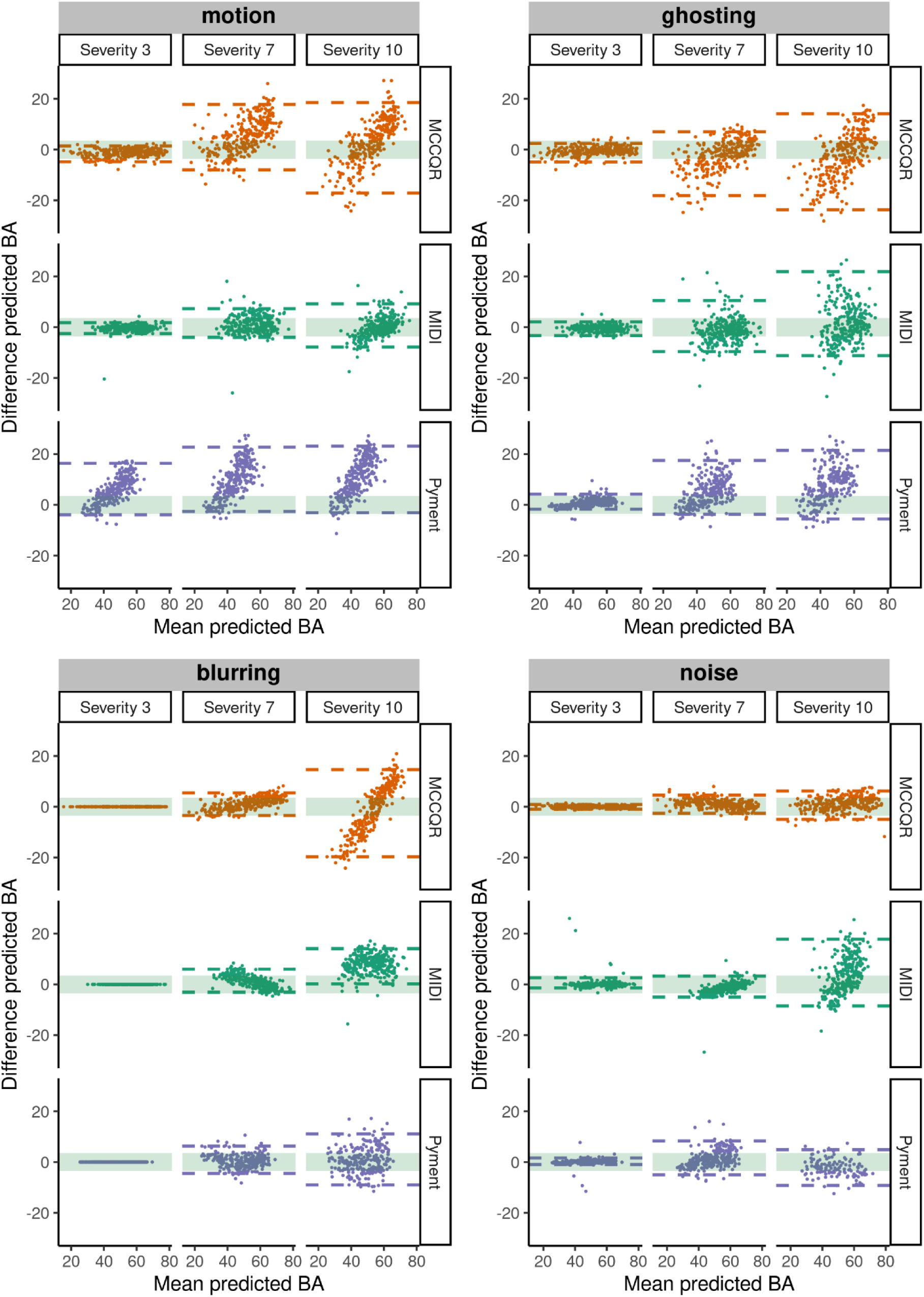
Bland–Altman plots comparing brain age predictions between artifact-free and degraded images (Severity 3, 7, and 10) across models (Pyment, MIDI, MCCQR) and artifact types. Dashed lines indicate the limits of agreement. The shaded green band marks the maximal allowable difference (MAD = ±3.55 years), representing a clinically relevant threshold for brain age acceleration (Hanson et al., 2024).

**Suppl. Figure 3.**
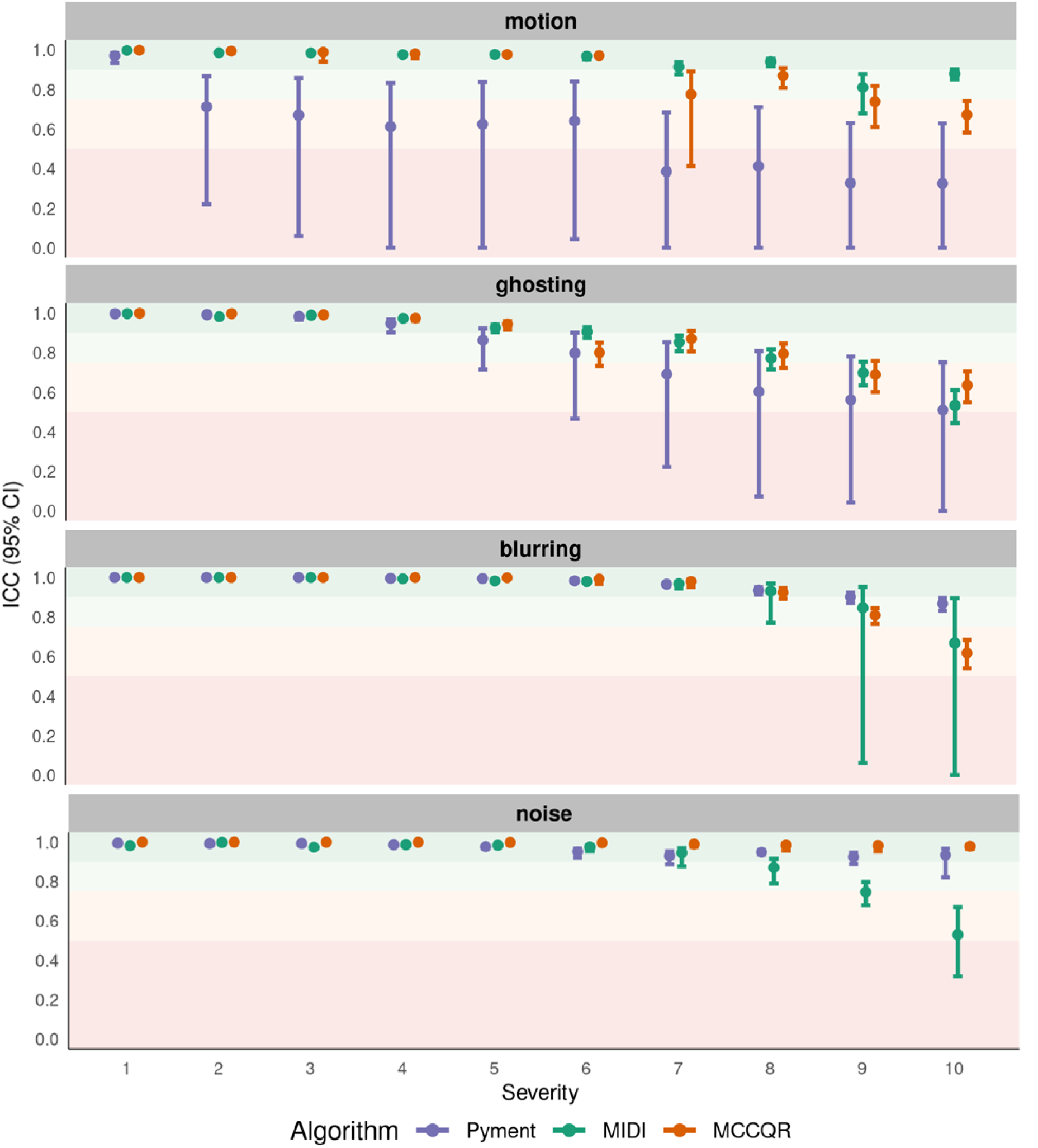
Impact of artifact severity on prediction stability after age-bias correction. Intraclass correlation coefficient (ICC) across severity levels, with shaded horizontal bands representing interpretive categories (Koo & Li, 2016): poor (<0.50, red), moderate (0.50–0.75, orange), good (0.75–0.90, light green), and excellent (>0.90, darker green). Points indicate estimates at each severity level, and vertical bars show 95% confidence intervals.

## Notes

### Competing Interest Statement

Richard Joules reports financial support was provided by IXICO plc. Richard Joules reports a relationship with IXICO plc that includes: employment.
Oscar Pena-Nogales reports a relationship with QMENTA Inc. that includes: employment.
Paulo Reis Rodrigues reports financial support was provided by QMENTA Inc. Paulo Reis Rodrigues reports a relationship with QMENTA Inc. that includes: board membership, employment, and equity or stocks. Paulo Reis Rodrigues has patent #WO2020210826A1 pending to Mint Labs Inc d/b/a QMENTA Inc.
Frederik Barkhof reports a relationship with Combinostics that includes: consulting or advisory. Frederik Barkhof reports a relationship with IXICO plc that includes: consulting or advisory. Frederik Barkhof reports a relationship with Roche that includes: consulting or advisory. Frederik Barkhof reports a relationship with Scottish Brain Sciences that includes: consulting or advisory. Frederik Barkhof reports a relationship with EISAI that includes: board membership. Frederik Barkhof reports a relationship with Biogen that includes: board membership. Frederik Barkhof reports a relationship with Prothena that includes: board membership. Frederik Barkhof reports a relationship with Merck that includes: board membership. Frederik Barkhof reports a relationship with Queen Square Analytics that includes: equity or stocks. Frederik Barkhof reports a relationship with EPSRC that includes: funding grants. Frederik Barkhof reports a relationship with EU-JU (IMI) that includes: funding grants. Frederik Barkhof reports a relationship with NIHR-BRC that includes: funding grants. Frederik Barkhof reports a relationship with GEHC that includes: funding grants. Frederik Barkhof reports a relationship with ADDI that includes: funding grants.
Henk Mutsaerts reports a relationship with TheriniBio that includes: consulting or advisory.
If there are other authors, they declare that they have no known competing financial interests or personal relationships that could have appeared to influence the work reported in this paper.

### Funding Statement

This project was funded by Health~Holland, Top Sector Life Sciences & Health (TKI-PPP Amsterdam UMC 2011227).

### Author Declarations

The ABRIM study fell under the ethics approval "Image Human Cognition" (Commissie Mensgebonden Onderzoek Arnhem-Nijmegen, 2014/288), and was approved by the Social Sciences Institutional Review Board of the Radboud University (ECSW 2017-3001-46).

